# Age-dependent somatic expansion of the *ATXN3* CAG repeat in the blood and buccal cell DNA of individuals with spinocerebellar ataxia type 3

**DOI:** 10.1101/2024.03.13.24303081

**Authors:** Ahmed M. Sidky, Ana Rosa Vieira Melo, Teresa T. Kay, Mafalda Raposo, Manuela Lima, Darren G. Monckton

**Author notes:** Surgery Brain Research Institute, J219, 5841 S. Maryland Avenue, Chicago, IL, 60637, USA.

## Abstract

Spinocerebellar ataxia type 3 (SCA3), a currently untreatable disorder, is caused by the expansion of a genetically unstable polyglutamine-encoding complex CAG repeat in the *ATXN3* gene. Longer alleles are generally associated with earlier onset and frequent intergenerational expansions mediate the anticipation observed in this disorder. Somatic expansion of the repeat has also been implicated in disease progression and slowing the rate of somatic expansion in patients has recently been proposed as a therapeutic strategy. Here, we utilised high-throughput ultra-deep MiSeq amplicon sequencing of the *ATXN3* repeat to precisely define the number of repeats, the exact sequence structure, the phased genotype of an adjacent single nucleotide polymorphism and to accurately quantify somatic expansion in blood and buccal cell DNA samples of a cohort of individuals with SCA3 from the Azores islands (Portugal). We revealed systematic mis-sizing of the *ATXN3* repeat and high levels of inaccuracy of the traditional fragment length analysis approach that have important implications for attempts to identify modifiers of clinical and molecular phenotypes, including genetic instability. Quantification of somatic expansion in blood DNA revealed the expected effects of age and CAG repeat length, although the effect of repeat length was surprisingly modest with much stronger associations with age at sampling. We also observed an association of the downstream rs12895357 single nucleotide polymorphism with the rate of somatic expansion, and a higher level of somatic expansion in buccal cell DNA compared to blood. Although the levels of somatic expansion are much lower per repeat unit, the average level of somatic expansion at the *ATXN3* locus in SCA3 patients is much higher than is typically observed at the *HTT* locus in Huntington disease patients. These data suggest that the *ATXN3* locus in SCA3 patients in blood or buccal cell DNA might serve as a good biomarker for clinical trials testing suppressors of somatic expansion with peripheral exposure.

## Introduction

Spinocerebellar ataxia type 3 (SCA3) (also known as Machado-Joseph disease (MJD)) is a rare autosomal dominantly inherited neurodegenerative disorder with a highly variable age at onset (Scott *et al*. 2020; Klockgether *et al*. 2019). SCA3-affected individuals show, in variable degrees, progressive gait ataxia, nystagmus, dysarthria, dystonia, distal muscle atrophy, ophthalmoplegia and fasciculations (Scott *et al*. 2020). SCA3 was first clinically reported in two Portuguese Azorean families that had emigrated to Massachusetts, USA (Nakano *et al*. 1972; Woods and Schaumburg 1972). SCA3 remains more frequent in Portugal, particularly in the Azores islands, with an overall prevalence of ∼ 1/2,500 and a remarkable 1/158 on Flores Island (Lima *et al*. 2023). Although the worldwide prevalence of SCA3 is much lower at ∼ 1/100,000, SCA3 is nonetheless considered the most common SCA accounting for about 20 to 50% of SCA families (Klockgether *et al*. 2019).

SCA3 is caused by the expansion of a polyglutamine-encoding CAG repeat in exon 10 of the *ATXN3* gene (NM_004993.6) (Kawaguchi *et al*. 1994). SCA3 is thus one of a group of disorders associated with the expansion of a glutamine-encoding CAG repeat, including the SCAs 1, 2, 7 and 17, DRPLA and Huntington disease (HD), that share a similar genetic mechanism and are assumed to also share a common toxic gain of function of the polyglutamine domain in the resultant protein (Bunting *et al*. 2022). The polyglutamine disorders are themselves a subset of a larger group of repeat expansion disorders that share a similar genetic basis, albeit with a variety of downstream pathological processes (Depienne and Mandel 2021). The *ATXN3* polyglutamine-encoding CAG repeat varies from ∼ 13 to ∼ 44 in the general population (Gardiner *et al*. 2019), and from ∼ 55 to ∼ 90 in individuals affected by SCA3 (de Mattos *et al*. 2019; Akcimen *et al*. 2020). As with other repeat expansion disorders (Depienne and Mandel 2021), measured repeat length is inversely correlated to age at onset accounting for ∼ 55 to 60% of the variation in disease severity (de Mattos *et al*. 2019; Akcimen *et al*. 2020). The *ATXN3* repeat is complex with the reference sequence (NM_004993.6) incorporating two synonymous glutamine-encoding CAA variants at positions three and six, and a non-synonymous lysine-encoding AAG variant at position four *i.e.,* (CAG)_2_(CAA)_1_(AAG)_1_(CAG)_1_(CAA)_1_(CAG)_x_ (Kawaguchi *et al*. 1994; Igarashi *et al*. 1996) (Figure 1). Direct Sanger sequencing of the *ATXN3* repeat in a small number of unaffected individuals (*n* = 36) and eleven patients from Japan (Kawaguchi *et al*. 1994) identified the presence of a sequence variant in which the CAA codon at position six is lost (see Figure 1C). This loss-of-CAA variant is relatively common (∼ 12%) on non-disease associated chromosomes in the Japanese population, but appears less common (∼ 2 to 5%) in other populations tested indirectly using allele-specific hybridisation technology (Kawaguchi *et al*. 1994; Igarashi *et al*. 1996). As far as we are aware, neither the loss-of-CAA variant, or any other sequence variants have been identified within the *ATXN3* repeat on disease-causing chromosomes, albeit only very few disease-associated alleles appear to have been sequenced in their entirety. The repeat tract is also followed immediately by a non-synonymous G/C single nucleotide polymorphism (SNP) in a glycine codon (rs12895357, NM_004993.6(ATXN3):c.916G>C (p.Gly306Arg)) (Figure 1) (Kawaguchi *et al*. 1994; Igarashi *et al*. 1996). The non-reference arginine-encoding C-allele of rs12895357 is very common (Auton *et al*. 2015) and is the major allele on non-disease associated chromosomes worldwide (Igarashi *et al*. 1996). Likewise, most *ATXN3* disease associated chromosomes are in phase with the C-allele, including all those reported in east Asian populations, but a subset of disease associated chromosomes in populations of European ancestry are in phase with the less common glycine-encoding G-allele (Igarashi *et al*. 1996).

**Figure 1.**
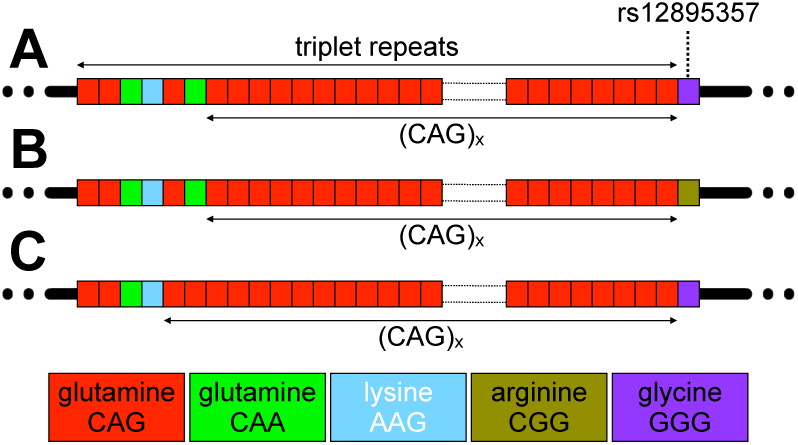
Allelic variation at the *ATXN3* triplet repeat locus. **A)** The schematic diagram shows the typical reference allele structure of the triplet repeat region in *ATXN3*, including the longest stretch of pure CAG repeats ((CAG)x). **B)** The non-reference arginine-encoding C-allele at the rs12895357 SNP. **C)** An atypical allele with loss of the CAA glutamine-encoding codon at repeat position six. The figure shows schematic representations of the alleles observed. Repeat codons are depicted: CAG glutamine codons as red boxes; CAA glutamine codons as green boxes; AAG lysine codons as sky blue boxes, CGG arginine codons as asparagus boxes; and GGG glycine codons as purple boxes.

One of the most characteristic features of the repeat expansion disorders is that the expanded disease associated alleles are intergenerationally unstable, with a bias toward expansion, explaining the anticipation observed in these disorders (Depienne and Mandel 2021). SCA3 is no exception, and intergenerational expansions are observed in ∼ 55% of transmissions, explaining the anticipation observed (Maciel *et al*. 1995; Sasaki *et al*. 1995; Durr *et al*. 1996; Igarashi *et al*. 1996; Souza *et al*. 2016). Interestingly, the genotype of the rs12895357 SNP has been implicated in modulating the degree of intergenerational expansion (Igarashi *et al*. 1996; Takiyama *et al*. 1997; Maciel *et al*. 1999). Heterozygotes carrying the C-allele in phase with the expanded disease-associated allele reportedly show higher levels of intergenerational instability in families and in sperm DNA directly, relative to homozygotes for either the C- or G-alleles (Igarashi *et al*. 1996; Takiyama *et al*. 1997; Maciel *et al*. 1999).

In addition to germline instability, again as with many repeat expansion disorders (Depienne and Mandel 2021), several small studies have revealed disease-causing *ATXN3* expansions to be somatically unstable in various tissues, including blood and brain (Cancel *et al*. 1995; Lopes-Cendes *et al*. 1996; Tanaka *et al*. 1996; Ito *et al*. 1998; Maciel *et al*. 1997; Cancel *et al*. 1998; Hashida *et al*. 1997; Munoz *et al*. 2002).

Despite the presumed critical role of the polyglutamine domain in cellular pathology (Bunting *et al*. 2022), recent data in HD, in which the polyglutamine domain is encoded by a mix of CAG and CAA codons, have revealed that the length of the pure CAG tract is a better predictor of disease severity than the length of the glutamine-encoding CAG/CAA tract (Ciosi *et al*. 2019; Genetic Modifiers of Huntington’s Disease Consortium *et al*. 2019; Wright *et al*. 2019). This observation is assumed to be primarily driven by the pre-eminent role of pure CAG length in driving somatic expansion (Ciosi *et al*. 2019), and the importance of somatic expansion in modulating disease onset (Ciosi *et al*. 2019; Genetic Modifiers of Huntington’s Disease Consortium *et al*. 2019; Hong *et al*. 2021). Indeed, in HD, after correction for CAG and age-effects, somatic expansion of the CAG repeat measured in blood, is inversely correlated with variation in age at onset (Ciosi *et al*. 2019). This interpretation of the mechanism by which pure CAG repeat length best predicts pathology, is further supported by the preponderance of DNA repair genes implicated in the somatic expansion process in animal models (Wheeler and Dion 2021) that have been revealed as modifiers of HD onset in genome-wide association studies (GWAS) (Genetic Modifiers of Huntington’s Disease Consortium *et al*. 2019; Hong *et al*. 2021).

The best practice guidelines for the molecular diagnosis of SCA3 is to PCR amplify across the repeat region, estimate fragment length by capillary gel electrophoresis and compare to known molecular weight standards, to estimate the number of repeats assuming the canonical repeat structure (Sequeiros *et al*. 2010a; Sequeiros *et al*. 2010b). Despite the fact that on average each extra *ATXN3* CAG repeat is associated with an approximately 2- to 3-year decrease in SCA3 age at onset (Akcimen *et al*. 2020; de Mattos *et al*. 2019), the guidelines allow for an error margin of +/- 3 repeats (Sequeiros *et al*. 2010a; Sequeiros *et al*. 2010b). Additionally, in a comparative study in which different protocols were used, only 17% of expanded alleles were sized the same between two independent laboratories, and ∼ 6% differed by more than +/- 3 repeats (Ramos *et al*. 2016). In addition to frequent discrepancies in absolute length between laboratories, fragment length analysis reveals no information on the known polymorphic rs12895357 SNP, or potentially disease-modifying sequence variants within or flanking the repeat. Previously, we utilised high-throughput ultra-deep MiSeq amplicon sequencing to precisely genotype the *HTT* CAG repeat, revealing the critical role for repeat-sequence in driving disease severity, and accurately quantifying somatic expansion in the blood DNA of individuals with HD (Ciosi *et al*. 2019). It was our hypothesis that high throughput ultra-deep MiSeq amplicon sequencing of the *ATXN3* repeat might provide similar insights in individuals with SCA3.

## Results

### High-throughput ultra-deep MiSeq sequencing and genotyping of the *ATXN3* repeat in individuals inheriting the SCA3 mutation

In order to assess sequence diversity at the *ATXN3* repeat in the SCA3 population, we adapted the high-throughput ultra-deep MiSeq sequencing assay we developed for analysing the HD-associated *HTT* CAG repeat (Ciosi *et al*. 2018; Ciosi *et al*. 2019) to the *ATXN3* locus. Briefly, the *ATXN3* repeat was PCR amplified using locus-specific primers incorporating MiSeq sequencing adaptors and spacers and sample-specific barcodes (allowing up to 384 barcode combinations). Amplicons were pooled, purified, and sequenced using MiSeq with 400 nt forward reads and 200 nt reverse reads, with an average read depth of ∼ 24,000 aligned reads per sample. We used this approach to attempt to sequence the *ATXN3* repeat from the blood DNA of 142 individuals inheriting SCA3-associated expansions belonging to the extensively studied Azorean cohort (Lima *et al*. 2023). In addition, we also sequenced the *ATXN3* repeat in paired buccal cell swab DNA samples in a subset of eleven individuals. Individuals were genotyped by aligning the forward reads to custom *ATXN3* reference sequences comprising a variable number of pure CAG repeats (0 to 100) and G or C at the site of the rs12895357 SNP which is located at the very first base after the CAG repeat region (Figure 1). This allowed simple phasing of rs12895357 to the relevant allele in heterozygotes, and provided a read length distribution for each participant (see Figure S1).

Traditionally, SCA3 individuals are diagnosed by PCR amplification of the *ATXN3* repeat from blood DNA and the genotype defined as the number of repeats estimated in the modal peak on fragment length analysis (Sequeiros *et al*. 2010a; Sequeiros *et al*. 2010b). The read-depth for the shorter non-disease-associated alleles was generally very high (∼ 17,500 aligned phased reads per sample), and in the vast majority of samples (*n* = 139) a clear modal allele was apparent (Figure S2). However, there were two samples with a phased aligned read depth for the non-disease associated allele of << 50 reads, and, in these cases, it was not possible to confidently define a modal repeat length. In addition, the modal repeat length observed in the MiSeq read length distributions for the shorter non-disease-associated alleles was the same in ten of the paired blood and buccal swab DNA samples. Most of the shorter non-disease-associated alleles (∼ 74%, 98/137) were also genotyped as the same modal length using MiSeq and fragment length analysis (*r*^2^ = 0.989, *p* < 2 x 10^-16^, *n* = 137 (estimation of repeat number by fragment length analysis was not available for two samples, Figure S3A). However, ∼ 24% (33/137) of these non-disease-associated alleles were estimated at one repeat smaller by fragment length analysis, two at two repeats smaller (∼ 1.5%), and one at one repeat bigger (∼ 0.7%). These differences were more pronounced for larger non-expanded alleles (*r*^2^ = 0.095, *p* = 0.0001, Figure S3B) with ∼ 94% (29/31) of 14 repeat alleles sized correctly, but only 65% (13/20) of 27 repeat alleles sized correctly, by fragment length analysis.

As expected, the lower PCR efficiency associated with larger fragments resulted in a reduced read depth for the expanded allele (∼ 6,500 aligned phased reads per sample). In cases with aligned phased read depths below ∼ 300 reads, it was not possible to confidently define a modal repeat length. Thus, we were only able to define the modal repeat length for 119/142 individuals (see Figure S4). The modal repeat length observed in the MiSeq read length distributions for the expanded alleles was the same in all the paired blood and buccal swab DNA samples (9/11) for which there was sufficient read depth (> 300 reads for the expanded allele in both samples, Figure 2). Fragment length data was available for most of these individuals (98%, 117/119), but although there was a high correlation between the modal repeat lengths determined by MiSeq and fragment length analysis (*r*^2^ = 0.885, *p* < 2 x 10^-16^, Figure S3C), all of the longer disease-associated alleles were sized shorter by fragment length analysis. The majority were estimated at either or two (36%, 42/117) or three (∼ 35%, 41/117) repeats smaller by fragment length analysis, with a smaller number at one (∼ 2.6%, 3/117), four (∼ 10%, 12/117), five (∼ 11%, 13/117), or even six repeats shorter (∼ 5%, 6/117). These differences were not allele length-dependent (*r*^2^ ∼ 0, *p* = 0.35, Figure S3D).

**Figure 2.**
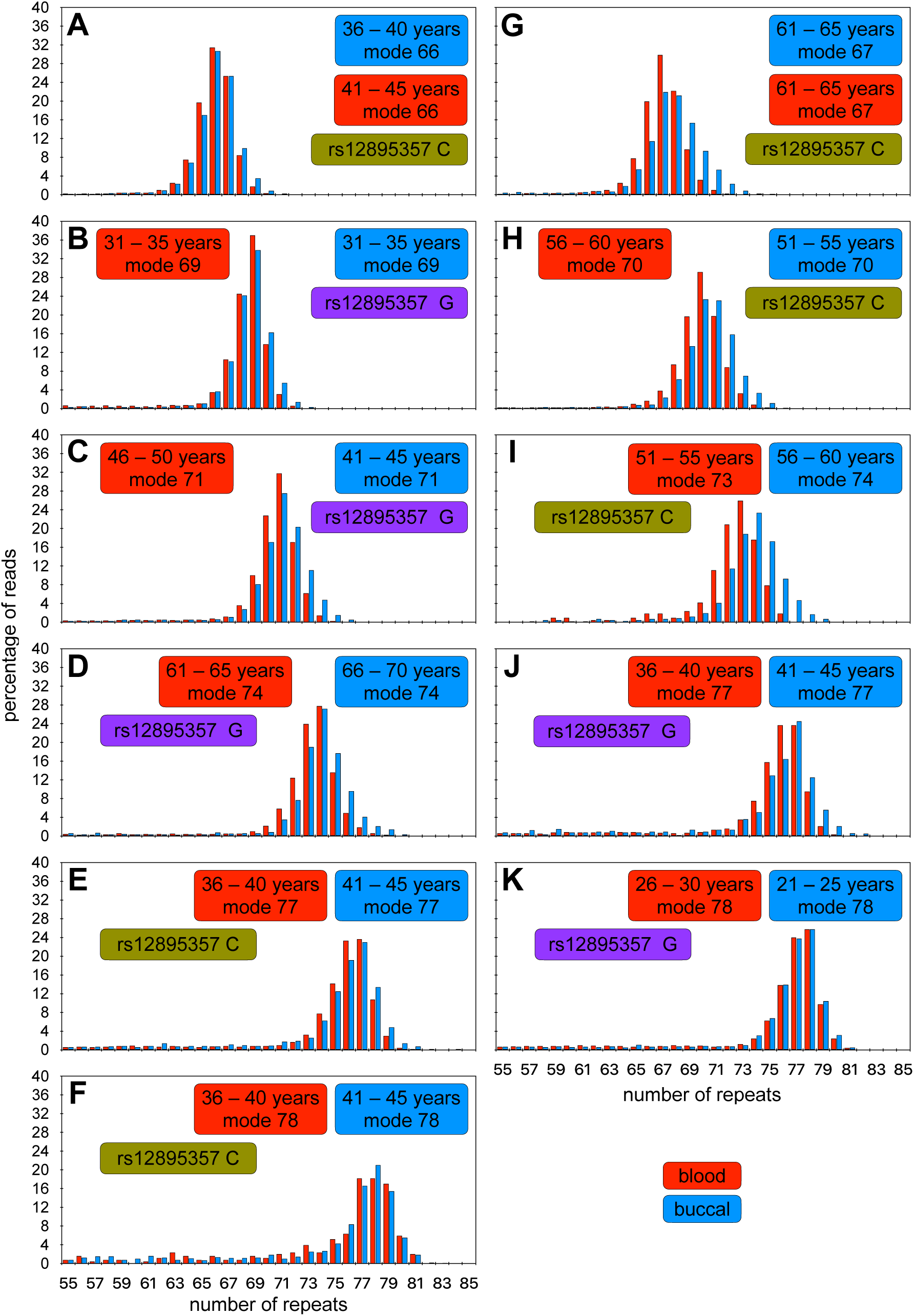
*ATXN3* triplet repeat read-length distributions for expanded disease-associated alleles in blood and buccal cell DNA. **A to K)** The histograms show read-length distributions for the expanded disease-associated *ATXN3* allele from both blood (red) and buccal (blue) cell DNA. MiSeq reads were aligned against references containing a variable number of CAG repeats and either the rs12895357 C-allele (asparagus) or G-allele (purple). For each participant the age at sampling and modal allele length observed for each tissue is indicated.

All the disease-associated expanded alleles (100%, 119/119), and the vast majority of non-disease associated alleles (∼99.25%, 134/135), were revealed to present with the typical allele structure matching the genomic reference sequence, varying only in the number of pure (CAG)_x_ repeats and in their genotype at rs12895357. Only one non-disease associated allele with loss of the second CAA glutamine-encoding repeat (see structure in Figure 1C) was observed in an allele with a total repeat length of 22 repeats and a pure (CAG)_x_ tract of 18 repeats with the rs12895357 G-allele. After correcting for close family relationships, we observed allele frequencies of 67% G (32/48, 95% confidence interval (CI) = 53 to 80%) and 32% C (15/48, 95% CI = 20 to 47%) at rs12895357 on non-disease associated alleles. These frequencies are consistent with those observed in European populations in the 1,000 genomes project (73:27%) (Auton *et al*. 2015) and as previously reported (Kawaguchi *et al*. 1994; Igarashi *et al*. 1996). There were very high levels of linkage disequilibrium with all 14 and 23 repeat alleles in phase with rs12895357-G (Figure S5A). Notably, we observed both rs12895357 alleles in phase with the larger disease-associated alleles: ∼ 28% G (13/46, 95% CI = 16 to 44%); and ∼ 72% C (33/46, 95% CI = 56 to 84%), consistent with at least two independent founder events for SCA3 in the Azores (Lima *et al*. 1998). In all cases we observed the same phase between rs12895357 and disease-associated alleles within families, and clustering between islands (Figure S5B/C). However, we did observe both rs12895357 alleles associated with expanded alleles in different families on one of the islands, consistent with at least two independent founder events for SCA3 on São Miguel (Figure S5B/C).

Interestingly, one individual presented without a detectable non-disease associated allele in either the blood or buccal cell swab DNA samples by both fragment length analysis and MiSeq. The expanded disease-associated allele(s) in this person were sequenced at very high depth (> 19,000 phased aligned reads) in both buccal swab and blood DNA, and both distributions looked very similar, with the same clear mode at 66 repeats (Figure 2A) phased to the rs12895357-C allele. We assume that this individual inherited two expanded disease associated alleles of 66 and 67 repeats. Given the inability to partition somatic variants to the two alleles, this individual has been excluded from the formal downstream somatic expansion and genotype-phenotype analyses (although see below).

### Somatic expansion of the *ATXN3* repeat

It is well known that the PCR amplification of short tandem repeats typically results in the generation of a tail of Taq polymerase slippage products that are shorter than the input molecule(s) (see (Shinde *et al*. 2003)). Previously, by sequencing single DNA molecules containing the *HTT* CAG repeat, it was confirmed that PCR slippage generated a high proportion of artefactual products smaller than the progenitor allele, but only a very low proportion of PCR products, larger than the progenitor allele (Ciosi *et al*. 2019). These data suggest the vast majority of expanded PCR products larger than the inherited progenitor allele in the bulk DNA sequencing data of the disease-causing alleles represent genuine somatic expansions (Ciosi *et al*. 2019).

Inspection of the read length distributions for the small non-disease-associated *ATXN3* alleles revealed a clear mode (Figure S2A), representing the inherited progenitor allele (size = N), and the expected tail of shorter repeat length-dependent Taq polymerase slippage products (N_-1_ N_-2_, N_-3_, *etc.,* Figure S2B). The absence of products slightly larger than the inherited non-disease-associated alleles (N_+1_, N_+2_, N_+3_ *etc.*), suggest that, as expected, these alleles are somatically very stable. In contrast, the expanded disease-associated alleles presented with much more diverse read-length distributions with many reads greater than the mode (Figure 2, S4A–W). These diverse distributions likely reflect the combined contribution of PCR Taq polymerase slippage artefacts, and genuine somatic expansions. We assume that in the majority of cases: the modal allele represents the inherited progenitor allele (size = N); the tail of shorter products (N_-1_, N_-2_, N_-3_, N_-4_ *etc.*) represents primarily Taq polymerase slippage products; and that the tail of products larger than the mode (N_+1_, N_+2_, N_+3_, N_+4_ *etc.*), represent primarily somatic expansions (Figure S4X). As we have previously done for similar analyses at the *HTT* locus (Ciosi *et al*. 2019), for those individuals with expanded allele read depth > 1,000 we thus quantified the degree of somatic expansion by calculating the somatic expansion ratio: (sum(N_+1_ to N_+10_))/N. We then performed a series of regression models to investigate the drivers of somatic expansion. Surprisingly there was no detectable main effect of repeat length on the somatic expansion ratio (*r*^2^ = 0.010, *p* = 0.15, Figure 3A), but there was a clear main effect of age at sampling (*r*^2^ = 0.33, *p* = 1 x 10^-8^, Figure 3B). The lack of a detectable main effect for repeat length in a univariate analysis most likely reflects, at least in part, the consequences of the intergenerational expansion bias that mediates genetic anticipation and a gross ascertainment bias in diagnosed families with a strong inverse correlation between age at sampling and repeat length (*r*^2^ = 0.29, *p* = 3 x 10^-9^, Figure S6) *i.e.,* individuals with larger repeats in the later generations of families present with an earlier age at onset and are sampled at an earlier age. Indeed, a multivariate model revealed highly significant effects for both age at sampling and repeat length, although there was no detectable interaction effect (*r*^2^ = 0.37, *p* = 2 x 10^-11^, *p_age_* = 6 x 10^-12^, *p_repeat_* = 0.008). Adding sex into the model revealed no detectable effect (*r*^2^ = 0.37, *p* = 1 x 10^-10^, *p_age_* = 7 x 10^-12^, *p_repeat_* = 0.009, *p_sex_* = 0.98). However, the addition of the phased rs12895357 SNP yielded a substantive improvement in the model (*r*^2^ = 0.43, *p* = 8 x 10^-13^, *p_age_* = 2 x 10^-12^, *p_repeat_* = 0.002, *p_rs12895357_* = 0.002). These data suggest that the rs12895357 C-allele is associated with higher rates of somatic expansion.

**Figure 3.**
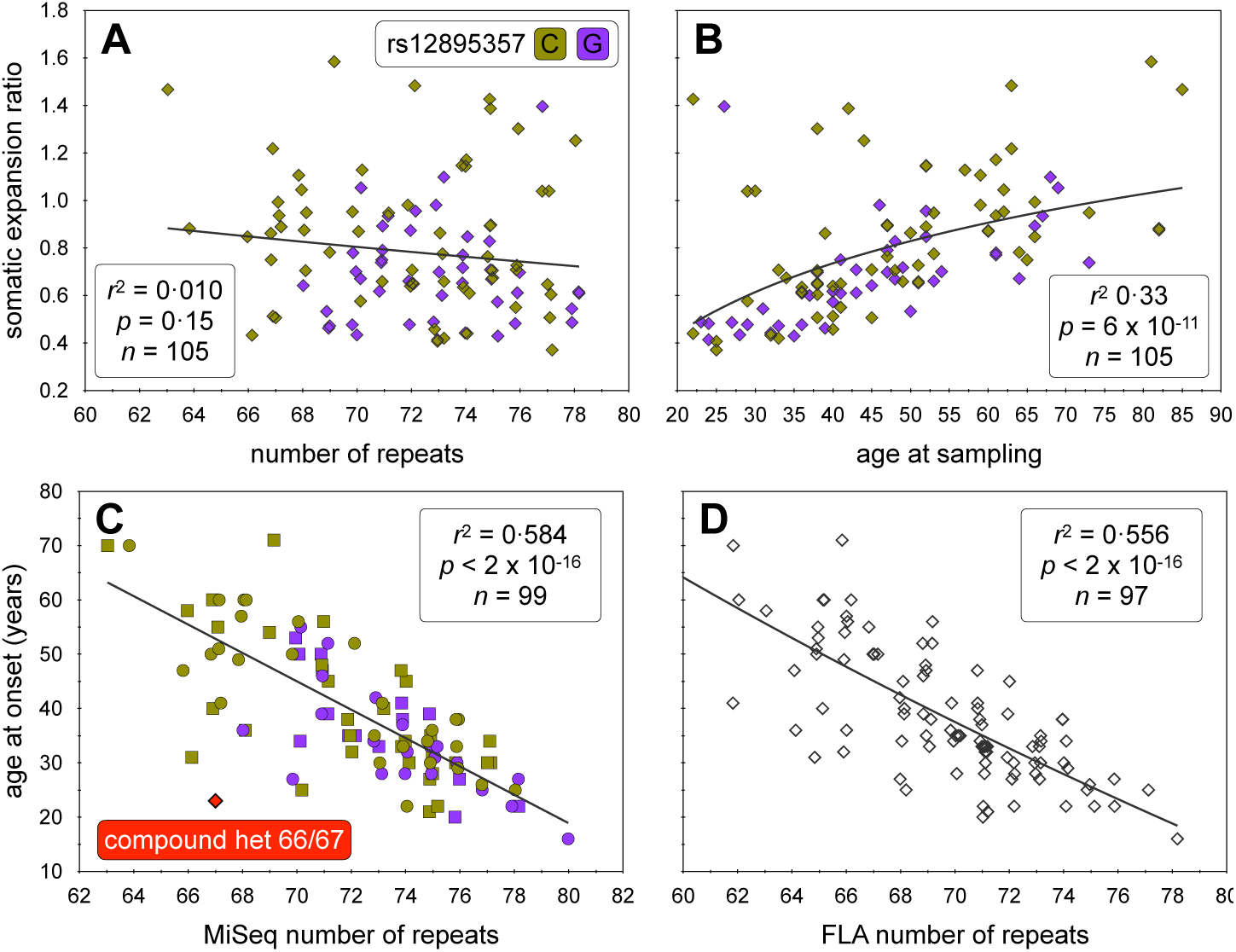
Modifiers of *ATXN3* triplet repeat blood DNA somatic expansion ratios and age at onset in SCA3. **A)** The scatterplot shows the ratio of somatic expansions of the expanded disease-associated *ATXN3* alleles measured in blood DNA dependent on the number of *ATXN3* repeats. **B)** The scatterplot shows the ratio of somatic expansions of the expanded disease-associated *ATXN3* alleles measured in blood DNA dependent on the age at sampling (years). **C)** The scatterplot shows the age at onset of SCA3 symptoms dependent on the number of *ATXN3* repeats as determined by MiSeq analysis in both females (circles) and males (squares). **D)** The scatterplot shows the age at onset of SCA3 symptoms dependent on the number of *ATXN3* repeats as determined by fragment length analysis. For each scatterplot the line of best fit (black line), adjusted coefficient of correlation squared (*r*^2^), *p*-value (*p*), and sample size (*n*) are indicated. For each participant rs12895357 genotype is also indicated: C-allele (asparagus) or G-allele (purple). Note, that to allow visualisation of overlapping points, random jitter (up to +/- 0.4 repeats) has been applied to the repeat numbers.

We also examined the degree of *ATXN3* somatic expansion in the DNA from eleven pairs of blood and buccal swab samples from the same individual (Figure 2). Despite the fact that some buccal swab samples were taken at an earlier age then the blood DNA samples, all individuals showed a higher somatic expansion ratio in the buccal swab DNA than in the blood DNA. Indeed, combining the buccal cell swab and blood somatic expansion ratio data into the multivariate model derived for blood and adding tissue as an additional independent variable, confirmed the rate of expansion is indeed greater in buccal swab DNA relative to blood cell DNA (*r*^2^ = 0.50, *p* = 2 x 10^-16^, *p_age_* = 1 x 10^-12^, *p_repeat_* = 0.02, *p_rs12895357_* = 0.001, *p_tissue_* = 6 x 10^-7^).

### Genotype-phenotype associations

As expected, the MiSeq modal allele length was inversely correlated with age at onset (*r*^2^ = 0.584, *p* < 2 x 10^-16^, *n* = 99, Figure 3C). The modal allele length determined by fragment length analysis was slightly worse at predicting age at onset (*r*^2^ = 0.556, *p* < 2 x 10^-16^, *n* = 97, Figure 3D). Neither sex, phased rs12895357 genotype, or residual variation in somatic expansion ratio in blood (corrected for repeat length, age and phased rs12895357 genotype) had any detectable association with age at onset (*r*^2^ = 0.575, *p* = 3 x 10^-15^, *p_repeat_* = 3 x 10^-16^, *p_sex_* = 0.492, *p_rs12895357_* = 0.242, *p_ResidualSER_* = 0.542, *n* = 99). Although not included in these formal genotype-phenotype analyses, it is apparent that the compound heterozygote inheriting two disease associated expansions of 66 and 67 repeats displays the largest deviation from the line of best fit with an age at onset ∼ 30 years earlier than might be expected (Figure 3C). Indeed, inclusion of this participant in the age at onset analyses using their largest allele (67 repeats) revealed participant zygosity to be a highly significant modifier (*r*^2^ = 0.587, *p* < 2 x 10^-16^, *p_zygosity_* = 0.0003, *n* = 100) with an effect size of -29.8 years for compound heterozygosity, consistent with previous observations of more severe SCA3 symptoms in individuals homozygous for *ATXN3* expansions (Lysenko *et al*. 2010).

### Intergenerational transmissions

In this cohort we were able to successfully determine the difference in modal allele length in 32 intergenerational transmissions (17 maternal, 15 paternal) (Figure S7). Intergenerational transmissions were significantly biased toward expansion (mean length change = +0.94 repeats, *p* = 0.012), as expected. Not surprisingly, given the very small sample size, we were unable to detect significant expected effects of sex of parent or repeat length on the mean length of intergenerational transmissions (*p* > 0.05, data not shown). Similarly, we could detect no effect of parental rs12895357 genotype, either on the expanded allele chromosome, the non-disease associated allele, or parental zygosity (*p* > 0.05, data not shown). However, we did note that there appeared to be greater variance in male transmissions (variance = 7.4 repeats, range = -3 to +7 repeats) relative to female transmissions (variance = 1.1 repeats, range = -1 to +2 repeats) (Figure S7B). This difference in variance between male and female transmission was highly significant (*F*-test, ratio of variances = 0.149, (95% CI = 0.051 to 0.421, *p* = 0.0005).

## Discussion

In this study we have used high-throughput ultra-deep MiSeq sequencing to successfully genotype the *ATXN3* CAG repeat in a large cohort of participants with SCA3 from the Azores. In addition to determining the modal repeat length, we were also able to determine the precise repeat structure, genotype and phase the rs12895357 SNP, and quantify the degree of somatic expansion for the disease-associated expanded alleles. Notably, we revealed a systematic mis-sizing of the expanded disease-associated allele with the traditional fragment length analysis consistently under-sizing the repeat length by two to three repeats (Figure 2C/D). More concerningly, some differences were as large as six repeats, and even in the best-case scenario assuming a systematic two repeat error, relative repeat sizes determined by fragment length analysis differed in 64% of cases (75/117). Whilst these differences have no impact on the ability to provide a yes/no diagnosis to patients, and have only a modest impact on observed genotype-phenotype relationships, they may have a more substantive impact on research studies. For instance, the primary phenotype used in SCA3 genetic modifier studies is residual age at onset *i.e.,* variation in age at onset not predicted by the repeat length of the expanded disease associated allele gene (de Mattos *et al*. 2019; Mergener *et al*. 2020; Akcimen *et al*. 2020; Raposo *et al*. 2022). Although such studies have started to reveal insights into pathways that may modulate SCA3, to date no genome-wide significant modifier variants have yet been defined. The two major factors mediating statistical power in genome-wide association studies are the robustness of the phenotype and the sample size. Whilst in general phenotypic robustness can be traded-off against sample size, in rare diseases such as SCA3, sample sizes are inevitably going to be rate limiting. Thus, there is a major impetus to increase the robustness of the phenotypes. Although residual variation in age at onset calculated using the MiSeq data and that calculated using the fragment length analysis data are highly corelated (*r*^2^ = 0.88, *n* = 97, *p* < 2 x 10^-16^), they are not the same and residual variation in age at onset calculated using MiSeq data should increase power in future genome-wide association studies. Even more substantively, mis-sizing repeat length by fragment length analysis has an even great impact on sizing intergenerational transmissions with only 20% of intergenerational changes sized the same (*r*^2^ = 0.52, *n* = 31, *p* = 3 x 10^-6^, Figure S7D, E). Such errors in sizing intergenerational transmissions have obvious implications for their prognostic utility, and in efforts to identify their biological determinants and genetic modifiers.

In HD, the HTT polyglutamine domain is encoded by a mix of CAG and CAA codons that exist in a variety of combinations in both non-disease, and disease associated expand alleles. Critically, in HD the length of the pure CAG tract is a better predictor of disease severity than the length of the glutamine-encoding CAG/CAA tract (Ciosi *et al*. 2019; Genetic Modifiers of Huntington’s Disease Consortium *et al*. 2019; Wright *et al*. 2019). Here, we revealed only one non-disease associated *ATXN3* allele with loss of the second CAA glutamine-encoding repeat (see structure in Figure 1C) that differed from the typical reference sequence. This loss-of-CAA variant is relatively common (∼ 12%) on non-disease associated chromosomes in the Japanese population (Kawaguchi *et al*. 1994; Igarashi *et al*. 1996), but is clearly much less common in the Azorean population (∼ 1%, 1/135 alleles), consistent with estimates from an allele-specific hybridisation assay (Igarashi *et al*. 1996). Likewise, previous analysis of expanded disease-associated alleles in the Azorean SCA3 population using an allele-specific hybridisation assay did not detect this loss-of-CAA variant (Igarashi *et al*. 1996). Here we have confirmed this observation, and revealed that no other *ATXN3* repeat sequence variants are present in expanded disease-associated alleles in the Azorean SCA3 population. Thus, the relative importance of pure CAG length versus encoded-polyglutamine repeat length in SCA3 cannot be currently assessed. However, the availability of a high-throughput MiSeq assay should increase the feasibility of screening additional more ethnically diverse SCA3 cohorts for potentially very insightful repeat sequence variants.

The greater prognostic value of pure CAG versus encoded-polyglutamine length in HD is assumed to be principally driven by the primary role of pure CAG length in influencing somatic expansion (Ciosi *et al*. 2019), and the importance of somatic expansion in modulating disease onset (Ciosi *et al*. 2019; Genetic Modifiers of Huntington’s Disease Consortium *et al*. 2019; Hong *et al*. 2021). Here, we utilised our MiSeq assay to quantify somatic repeat dynamics of the *ATXN3* repeat of the disease-associated expanded allele in the blood DNA of the Azorean SCA3 cohort. Somewhat surprisingly, in clear contrast to the *HTT* CAG repeat (Ciosi *et al*. 2019), we did not detect an obvious main effect of repeat number on the somatic expansion ratio (Figure S8A). However, again in contrast to *HTT*, we did observe a very strong main positive effect of age at sampling (Figure S8B). Nonetheless, as with *HTT*, in a multivariate model both age and repeat number were revealed as significant modifiers of the somatic expansion ratio. Whilst the lack of an obvious main positive effect for age at the *HTT* locus, or repeat length at the *ATXN3* locus, is likely at least partially attributable to the confounding effects of the age at sampling bias that characterises both disorders, the profound distinction between the two loci is noteworthy. One potential explanation is that most HD-associated *HTT* alleles are only just above the threshold at which somatic expansion in blood becomes detectable, and in a region where there is a clear exponential relationship between repeat length and somatic expansion. In contrast, the length of *ATXN3* alleles associated with SCA3 are much longer, and it is possible that the rate of change in somatic expansion with CAG repeat length is beyond the exponential phase. The lack of an obvious repeat length effect on somatic expansion appears to be mirrored in germline transmission, which again fail to reveal obvious parental allele length effects (Maciel *et al*. 1995; Durr *et al*. 1996; Martins *et al*. 2008; Souza *et al*. 2016) that are characteristic of HD and most other repeat expansion disorders (Depienne and Mandel 2021).

Another interesting comparison is the obvious difference in absolute somatic expansion ratios between *HTT* and *ATXN3* relative to repeat length. Even after correcting for pure CAG repeat length, it is clear that the *ATXN3* repeat is much more stable per CAG unit than the *HTT* locus (Figure S8A). These data are consistent with major inter-CAG-repeat-locus *cis*-acting modifiers, as has been previously observed for germline repeat dynamics (Brock *et al*. 1999; Nestor and Monckton 2011). Indeed, the data here confirm the speculation that the *ATXN3* repeat is substantively more stable in the soma than the *HTT* repeat (Nestor and Monckton 2011). This observation appears to be replicated in the brain where, using fluorescence-activated nuclear sorting, it has recently been demonstrated that although unstable in medium spiny neurons, the *ATXN3* repeat in SCA3 donors was much more stable per repeat unit than the *HTT* repeat in HD donors (Mätlik *et al*. 2024). These data provide a potential explanation for the major difference in relative polyglutamine toxicity between the two loci as exemplified by the differences in the disease-associated range between HD (typically 40 to 60 polyglutamine-encoding repeats) and SCA3 (typically 60 to 80 polyglutamine-encoding repeats) *i.e.,* that the intra-neuronal threshold for polyglutamine toxicity is higher than the inherited repeat length, and that individuals with SCA3 need to inherit a larger, but more slowly expanding, *ATXN3* allele to reach the cellular threshold at a similar age as an individual with HD inheriting a smaller, but more rapidly expanding, *HTT* allele (Nestor and Monckton 2011). A key role for somatic expansion in HD is also supported by the observation that residual variation in somatic expansion of the CAG repeat measured in blood (after correction for CAG and age-effects), is inversely correlated with variation in age at onset (Ciosi *et al*. 2019). We were not able to detect such an association in our SCA3 data. However, it should be noted that although highly significant, the effect size in HD was very modest, and was detected in a much larger cohort (*n* = 734) (Ciosi *et al*. 2019). Thus, analysis of a much larger cohort will be required to rigorously test this hypothesis in SCA3.

Somatic expansion is now widely viewed as an important therapeutic target in HD, and likely in other related neurological disorders caused by the expansion of short tandem repeats, and efforts are afoot to identify drugs that might suppress somatic expansion (Benn *et al*. 2021). An important component of a clinical trial of any such entity will be to demonstrate that the drug does indeed suppress somatic expansion *in vivo*. Whilst ideally this would be demonstrated in the affected neurons, this is clearly not practical. Thus, at least for drugs with peripheral exposure, there is interest in identifying biomarkers in accessible peripheral tissues.

Blood is the most obvious source of DNA for such analyses, but the levels of somatic expansion at the *HTT* locus in the majority of individuals with HD inheriting less than 50 repeats is very modest, and detecting changes over short time periods is likely to be technically challenging. Although the rate of expansion of the *ATXN3* repeat is lower per repeat unit than at *HTT*, the average inherited repeat length in SCA3 patients is much higher and the average rate of expansion is higher (Figure S8A). Despite the fact that SCA3 is much rarer than HD, the higher average rate of somatic expansion in SCA3, coupled with the lack of a clear main effect for repeat length, and the clear effect of age on somatic expansion of the *ATXN3* repeat in individuals with SCA3, suggest that SCA3 might present as an attractive disorder in which to establish proof-of-action in a clinical trial of a somatic expansion supressing drug with peripheral exposure. In addition, the demonstration here that the levels of somatic expansion were even higher in buccal cells as opposed to blood cells, suggest that *ATXN3* expansion in buccal cells may be an even more powerful biomarker. Further larger longitudinal studies will be required to better evaluate the biomarker potential of the *ATXN3* repeat in the SCA3 population.

Interestingly, we revealed here an apparent association of the rs12895357 C-allele with higher rates of somatic expansion in blood DNA. These data are consistent with the critical role of *cis*-acting elements in modulating inter-locus differences in expansion rates (Brock *et al*. 1999; Nestor and Monckton 2011), and suggest that naturally occurring *cis*-acting variants may also modulate intra-locus differences. These data may also relate to the previous observation that this variant may also be associated with altered rates of intergenerational expansion (Igarashi *et al*. 1996; Maciel *et al*. 1999; Takiyama *et al*. 1997). However, a simple *cis*-modifier effect of rs12895357 genotype on the intergenerational dynamics of the linked *ATXN3* repeat allele was not observed. Rather, rs12895357 heterozygotes carrying the C-allele phased with the expanded repeat appeared to have higher rates of instability compared to C/C or G/G rs12895357 homozygotes (Igarashi *et al*. 1996; Maciel *et al*. 1999; Takiyama *et al*. 1997). Such an effect could conceivably be mediated by inter-chromosomal interactions during meiosis, although this effect was not replicated in an independent analysis of sperm DNA analysis (Grewal *et al*. 1999). We observed no effect of rs12895357 on the intergenerational dynamics assessed in this study, but our sample size was very small (32 transmissions). Further larger studies will be required to better evaluate the effects of rs12895357 on both somatic and germinal *ATXN3* repeat dynamics.

In all of the analyses presented here, we have used the modal allele repeat length observed in the MiSeq distribution as the most objective estimate of the actual inherited allele length of disease-associated alleles. In many cases, the read length distributions present with a clear mode, a characteristic tail of shorter Taq polymerase slippage products typically extending to ∼ N_-5_, and a modest number of expansions typically extending to N_+3/4_ (*e.g.,* Figure S4C, E, J, K, M, S *etc.*). Such distributions are most commonly observed in samples from individuals sampled at a younger age (< 50 years), and with smaller modal alleles (< 74 repeats). In these cases, we are confident that the modal allele does indeed reflect the inherited progenitor allele length. However, in some individuals with longer alleles and/or sampled at greater ages, the repeat length distributions are more difficult to interpret. For instance, consider the distributions presented in Figure S4P and S4Q. Both have a very similar overall distribution over the same range of repeats, but the mode is one repeat different between them. Considering the distribution in Figure S4P, it is very credible that the mode of 75 represents the inherited progenitor allele and that the high peak at N_+1_ (76 repeats), reflects high levels of somatic expansion. Indeed, although we don’t consider it likely, it may even be possible that this allele is so unstable that the actual inherited allele was only 74 repeats and that even the modal peak at 75 repeats represents somatic expansion products. Conversely, it’s possible that the length dependent Taq polymerase slippage is so great that the mode at 75 actually represents primarily N_-1_ slippage products from a true inherited allele length of 76. Similar arguments can be made for the distribution presented in

Figure S4Q, with plausible inherited progenitor allele lengths from 74 to 76 repeats. Unfortunately, we currently have no objective method to distinguish these possibilities and derive the true inherited allele length or provide an alternative estimated progenitor allele length. This inability to objectively define the progenitor allele is the primary limitation of our study, as it will have been in all previous studies of expanded *ATXN3* repeat alleles. A critical imperative for future SCA3 studies is therefore to employ PCR-free long read sequencing approaches (*e.g.,* (Hafford-Tear *et al*. 2019)), or implement single molecule barcoding approaches to correct for PCR slippage (Woerner *et al*. 2021), in order to potentially identify the progenitor allele.

Here, we have established the utility of a high-throughput ultra-deep MiSeq sequencing assay to genotype the *ATXN3* repeat, expansions of which cause SCA3. In addition to accurately revealing repeat length, the assay also reveals the exact structure of the complex polyglutamine encoding repeat, phases the rs12895357 SNP and allows for quantification of somatic expansion. Our application of this assay to the Azorean SCA3 cohort has already revealed the limitations of the traditional fragment length analysis approach and yielded new insights into the key age- and tissue-dependence, and apparent modulatory effect of the rs12895357 SNP, on somatic expansion. These observations pave the way for the application of this technology to larger more diverse SCA3 cohorts to augment ongoing efforts to define the *cis*- and *trans*-acting modifiers of genetic instability and clinical phenotypes as therapeutic targets in SCA3.

## Materials and Methods

### Cohort

A group of 142 molecularly confirmed SCA3 affected subjects from the Azorean SCA3 cohort (Lima *et al*. 2023), were recruited for this study. Blood samples were used to determine the number of repeats in the *ATXN3* gene, define repeat structure, genotype the rs12895357 SNP, and quantify somatic expansions in blood genomic DNA of all participants, in addition to a subset of 11 paired buccal cell swab DNA samples. All participants were recruited with informed consent for genetic analyses and the study has been approved by the Ethical Board of the University of the Azores (Parecer 1/2020). Note that the Family IDs depicted in Figure S5 are utilised for research purposes only and are not known to anyone outside the research group.

### Illumina MiSeq sequencing of *ATXN3* CAG repeat

The previously reported locus-specific primers MJD52/MJD70 (Kawaguchi *et al*. 1994) were modified to incorporate Illumina MiSeq adapters sequences, Nextera XT indexes and sequencing diversity increasing spacers, allowing the sequencing of multiplexed 384 amplicons as previously described for the *HTT* locus (Ciosi *et al*. 2018; Ciosi *et al*. 2019). The *ATXN3* CAG repeat was amplified by PCR in a total volume of 10 μl containing 10 ng of genomic DNA, 1 μM of forward and reverse MiSeq primers, 1X Custom PCR master mix (Thermo Scientific, SM005) and 1 U of Taq polymerase (Sigma). PCR conditions were: an initial denaturation at 95°C for 5 minutes; followed by 25 cycles of denaturation (95°C for 1 minute); annealing (56°C for 1 minute); and extension (72°C for 1 minute); and a final extension at 72°C for 5 minutes. Library clean up and fragment size selection were carried out using 0.8x AMPure XP magnetic beads (Beckman Coulter) to remove primer dimers (Ciosi *et al*. 2018). DNA library concentration was quantified by Qubit™ fluorometry using the Qubit™ dsDNA HS Assay Kit (Thermo Scientific). To check the amplicon sizes, verify the absence of primer dimers and the presence of amplicons of interest, and accurately measure the molar concentration of the sequencing library, capillary electrophoresis was carried out on the 2100 Bioanalyzer using the Agilent High sensitivity DNA kit (https://www.agilent.com). The DNA library was sequenced and demultiplexed at the College for Medical Veterinary and Life Sciences Shared Research Facilities, a service provider at the University of Glasgow (https://www.polyomics.gla.ac.uk/ngs_omics.html). Amplicons were sequenced on the Illumina MiSeq platform using the MiSeq reagent v3 kit (Illumina). As a sequencing control, PhiX DNA was added to the library at 5% molar concentration. The *ATXN3* library was sequenced with 400 nt forward reads and 200 nt reverse reads in a 600 nt-cycle run. Demultiplexing of the MiSeq reads was carried out according to the Illumina i5 and i7 indices from Nextera XT index kit v2, then the demultiplexed reads were exported in FASTQ file format for the subsequent bioinformatic analyses (Ciosi *et al*. 2018).

### Bioinformatic analyses of MiSeq amplicon libraries

Custom *ATXN3* reference sequences were generated using the RefGeneratr tool (https://github.com/helloabunai/RefGeneratr) with variable CAG repeat numbers; (CAG)_1 – 100_(G/C). Bioinformatic analyses of Illumina MiSeq sequencing reads was performed via the open-source Galaxy interface (Afgan *et al*. 2016), hosted by the University of Glasgow (http://heighliner.cvr.gla.ac.uk). Bioinformatic pipelines were set, and Galaxy workflows were created to allow the simultaneous analysis of the MiSeq sequenced *ATXN3* library. Briefly, the workflow included demultiplexing the reads using the locus specific primer and adaptor trimming Cutadapt tool (Martin 2011), Followed by alignment of the trimmed MiSeq sequence reads to the *ATXN3* custom reference sequences using the Map-with-BWA_MEM tool (Li and Durbin 2009, 2010; Li *et al*. 2009). Following the alignment the generated BAM files which contain the mapped reads were then converted into SAM files by the BAM-to-SAM tool (Li *et al*. 2009). To remove reads with multiple alignments with the same quality score, additional filtering according to the mapping quality (MAPQ) score was applied so all alignments with MAPQ equal to zero were filtered out. Aligned filtered SAM files were downloaded and visualised using the Tablet genomic sequence viewer (Milne *et al*. 2013).

### Genetic and Statistical analyses

Statistical analyses were performed using R (version 4.3.2)(The R Core Team 2016) via RStudio (version 2023.12.1+402)(The RStudio Team 2016).All single and multiple linear regression models were carried out using the lm() function within the R stats package (The R Core Team 2016).

## Acknowledgments

The authors would like to thank all of the participants who donated valuable samples and data to allow this study. We gratefully thank Dr Aires Raposo for collaboration on blood collection. We further acknowledge the fundamental role of the Azorean Patients Association (Associação Atlântica de Apoio ao Doente de Machado-Joseph). The authors would also like to thank the DGM group for helpful advice during the conduct of the study. For the purpose of open access, the authors have applied a Creative Commons Attribution (CC BY) licence to any Author Accepted Manuscript version arising from this submission.

## Funding

The study was funded by a Newton Mosharafa PhD Scholarship award to AMS from the British Council and the Egyptian Ministry of Higher Education and Scientific Research. MR is supported by FCT (https://doi.org/10.54499/CEECIND/03018/2018/CP1556/CT0009). The Fundo Regional para a Ciência e Tecnologia (FRCT, Governo Regional dos Açores) has supported this work, under the PRO-SCIENTIA program.

## Competing interests

Within the last 36 months Professor Monckton has been a scientific consultant and/or received an honoraria/grants from AMO Pharma, Dyne, F. Hoffman-La Roche, LoQus23, MOMA Therapeutics, Novartis, Ono Pharmaceuticals, Pfizer Pharmaceuticals, Rgenta Therapeutics, Sanofi, Sarepta Therapeutics Inc, Script Biosciences, Triplet Therapeutics, and Vertex Pharmaceuticals. Professor Monckton also had research contracts with AMO Pharma and Vertex Pharmaceuticals. The other authors have declared no competing interests.

## Data Availability Statement

All data produced in the present study are available upon reasonable request to the authors.

---

**Figure S1.**
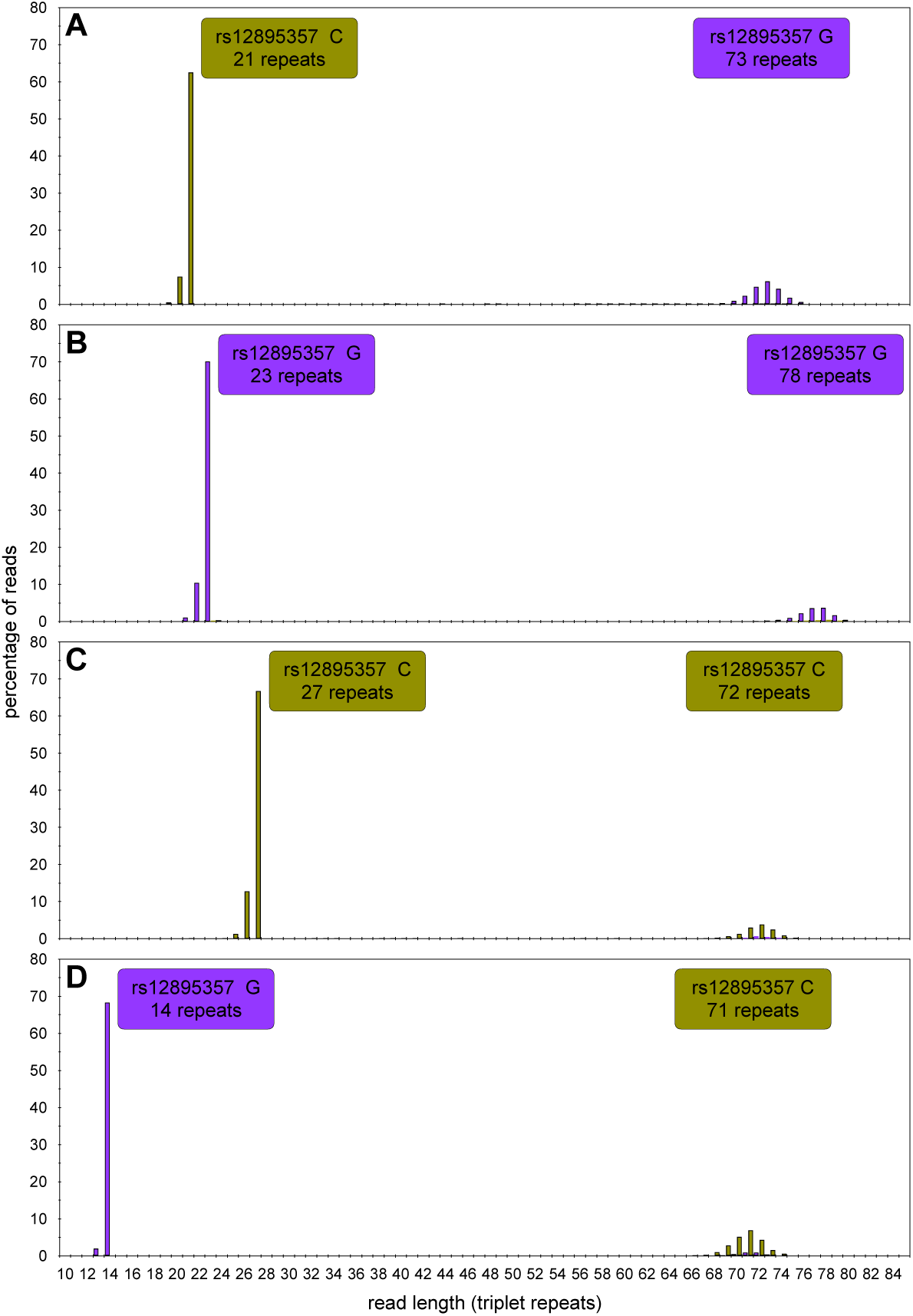
*ATXN3* triplet repeat rs12895357-specific read-length distributions. The histograms show the read-length distributions for four participants representing all four possible combinations of rs12895357 and expanded and non-expanded *ATXN3* triplet repeat alleles **(A – D)**. MiSeq reads were aligned against references containing a variable number of CAG repeats and either the rs12895357 C-allele (asparagus) or G-allele (purple). The two read length distributions allow the clear phasing of each triplet repeat allele to its cognate rs12895357 allele.

**Figure S2.**
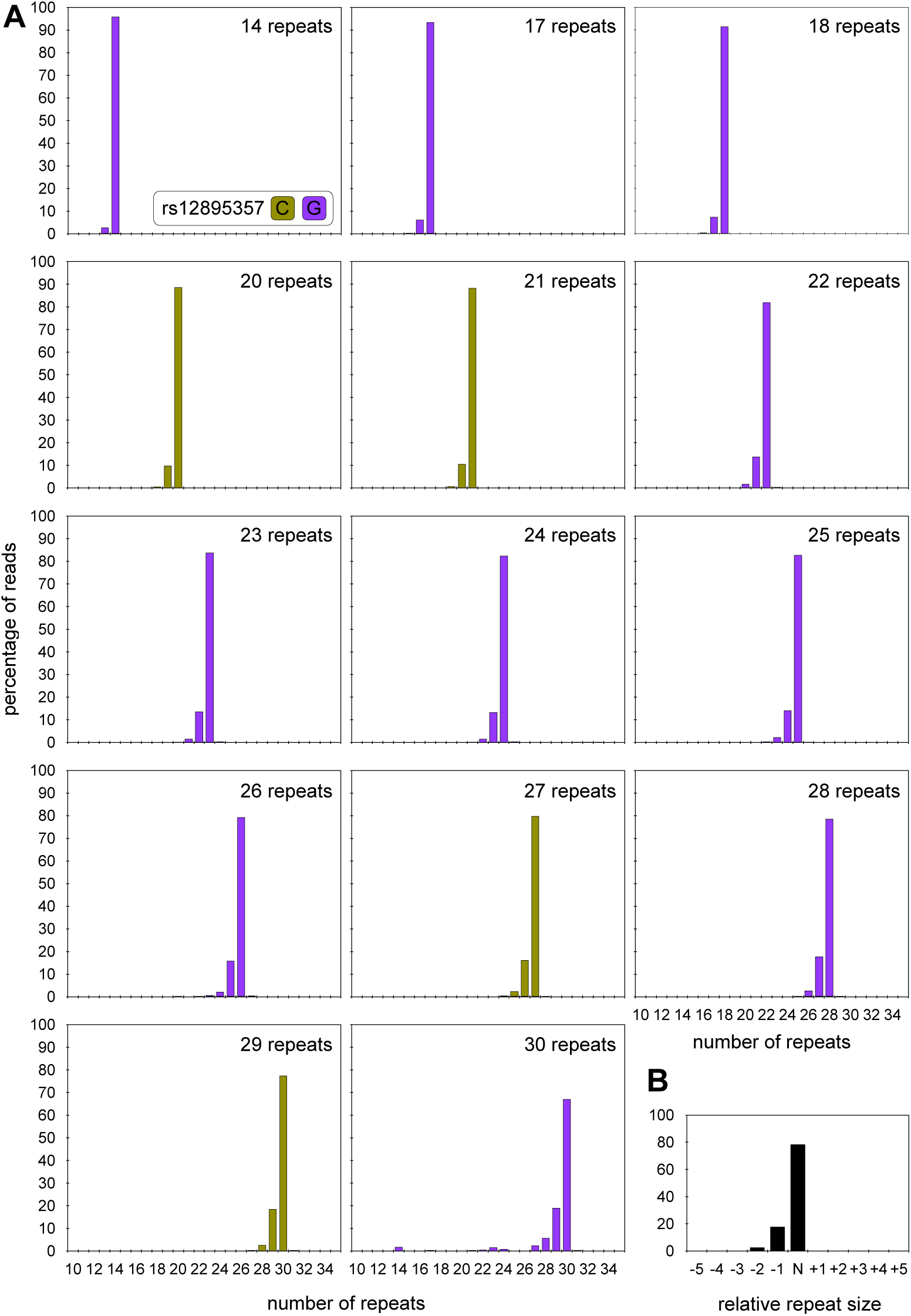
*ATXN3* triplet repeat read-length distributions for small non-disease associated alleles. **A)** The histograms show representative read-length distributions for the small non-disease associated *ATXN3* allele from participants covering the full range of such alleles. MiSeq reads were aligned against references containing a variable number of CAG repeats and either the rs12895357 C-allele (asparagus) or G-allele (purple). **B)** The histogram shows a representative read length distribution for the small non-disease associated *ATXN3* alleles. The major modal allele is defined as N, and with reads smaller than the mode indicated as N-1, N-2, N-3 *etc.,* and reads larger than the mode indicated as N+1, N+2 *etc*.

**Figure S3.**
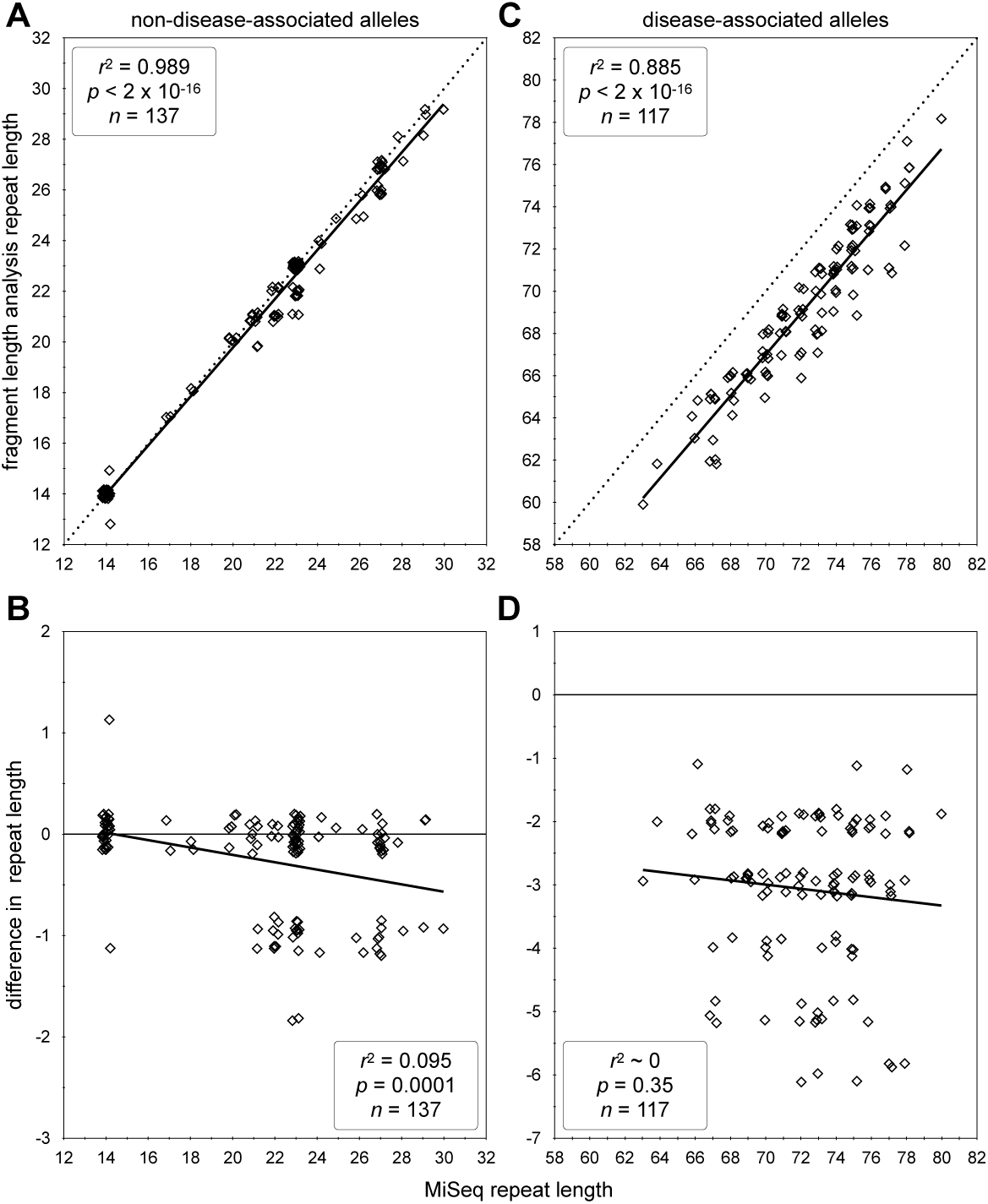
Modal allele length comparisons between *ATXN3* triplet repeat length determined by MiSeq and fragment length analysis. The scatterplots show MiSeq modal allele length plotted against absolute modal allele length as determined by fragment length analysis **(A, C)** or against the difference in modal allele length between MiSeq and fragment length analysis **(B, D)** for small non-disease associated alleles **(A, B)** and expanded disease-associated alleles **(C, D)**. In each case the line of best fit (dark black line), adjusted coefficient of correlation squared (*r*^2^), *p*-value (*p*) and sample size (*n*) are indicated. For **(A, C)** the expected 1:1 fit is also indicated with a dashed line. Note, that to allow visualisation of overlapping points, random jitter (up to +/- 0.4 repeats) has been applied to all data points.

**Figure S4.**
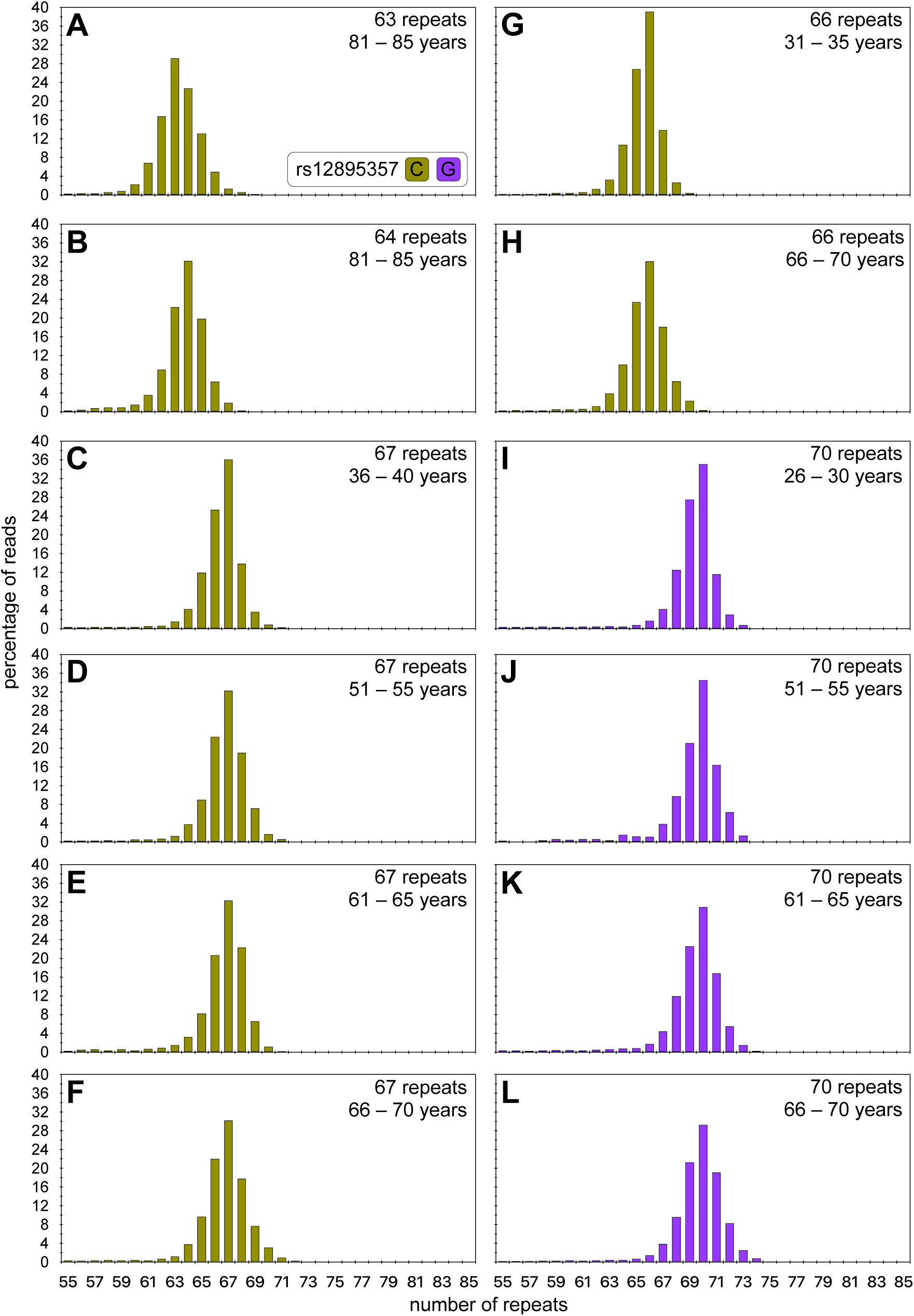
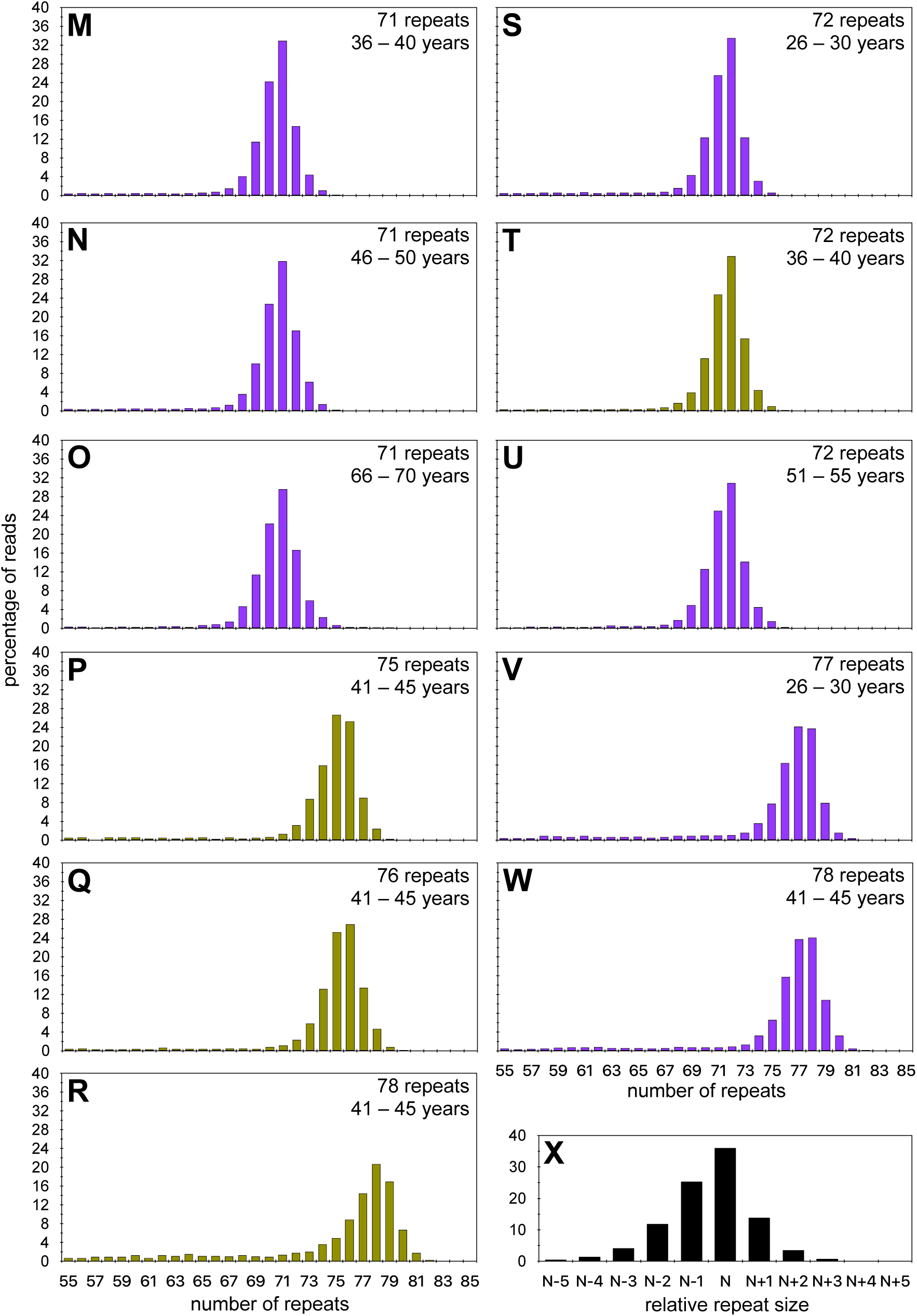
*ATXN3* triplet repeat read-length distributions for expanded disease-associated alleles. **A to W)** The histograms show representative read-length distributions for the expanded disease-associated *ATXN3* allele from participants covering the full range of such alleles. MiSeq reads were aligned against references containing a variable number of CAG repeats and either the rs12895357 C-allele (asparagus) or G-allele (purple). **X)** The histogram shows a representative read length distribution for expanded disease-associated *ATXN3* alleles. The major modal allele is defined as N, and with reads smaller than the mode indicated as N-1, N-2, N-3 *etc.,* and reads larger than the mode indicated as N+1, N+2, N+3 *etc*.

**Figure S5.**
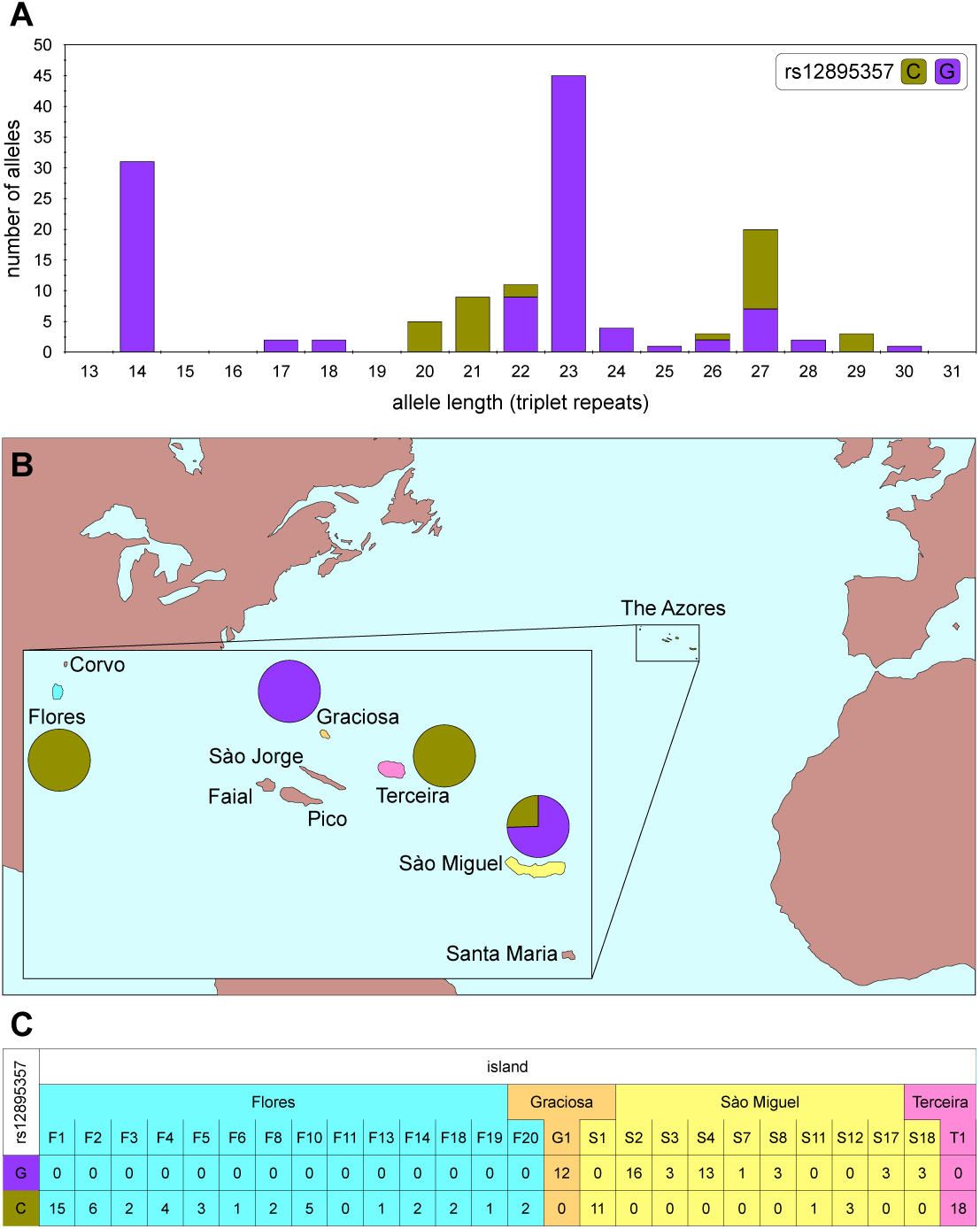
rs12895357 genotypes. **A)** The histogram shows rs12895357 *ATXN3* repeat length associations for the small non-disease-associated alleles. **B)** The map and pie charts show the geographical distribution for the rs12895357 alleles linked to expanded disease associated *ATXN3* repeat alleles in the Azores. **C)** The table shows the geographical and familial (F1, F2, G1, S1 *etc.*) distribution for the rs12895357 alleles linked to expanded disease-associated *ATXN3* repeat alleles in the Azores where each table entry indicates the number of family members with the specific genotype assayed.

**Figure S6.**
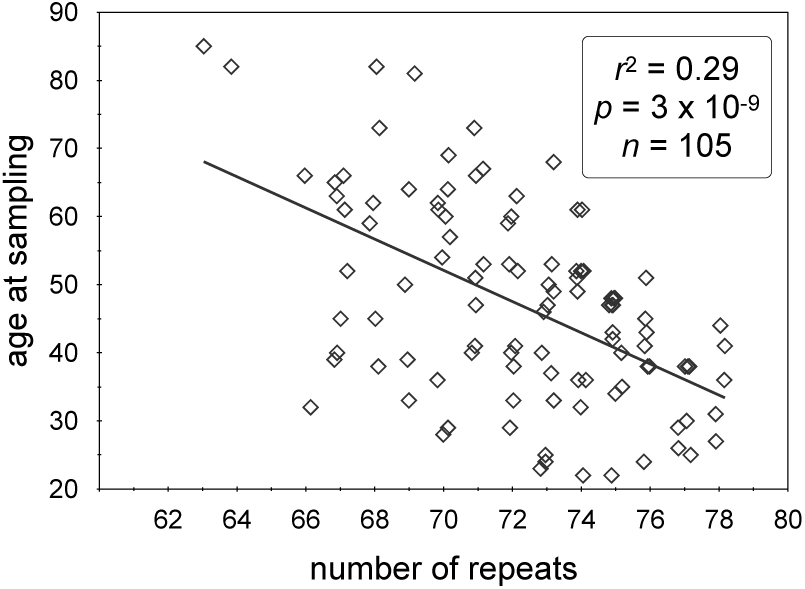
Age at sampling bias in SCA3. The scatterplot shows the inverse association between age at sampling of SCA3 and *ATXN3* expanded disease-associated allele repeat length in the Azorean SCA3 cohort. The line of best fit (black line), adjusted coefficient of correlation squared (*r*^2^), *p*-value (*p*), and sample size (*n*) are indicated. Note, that to allow visualisation of overlapping points, random jitter (up to +/- 0.4 repeats) has been applied to the number of repeats.

**Figure S7.**
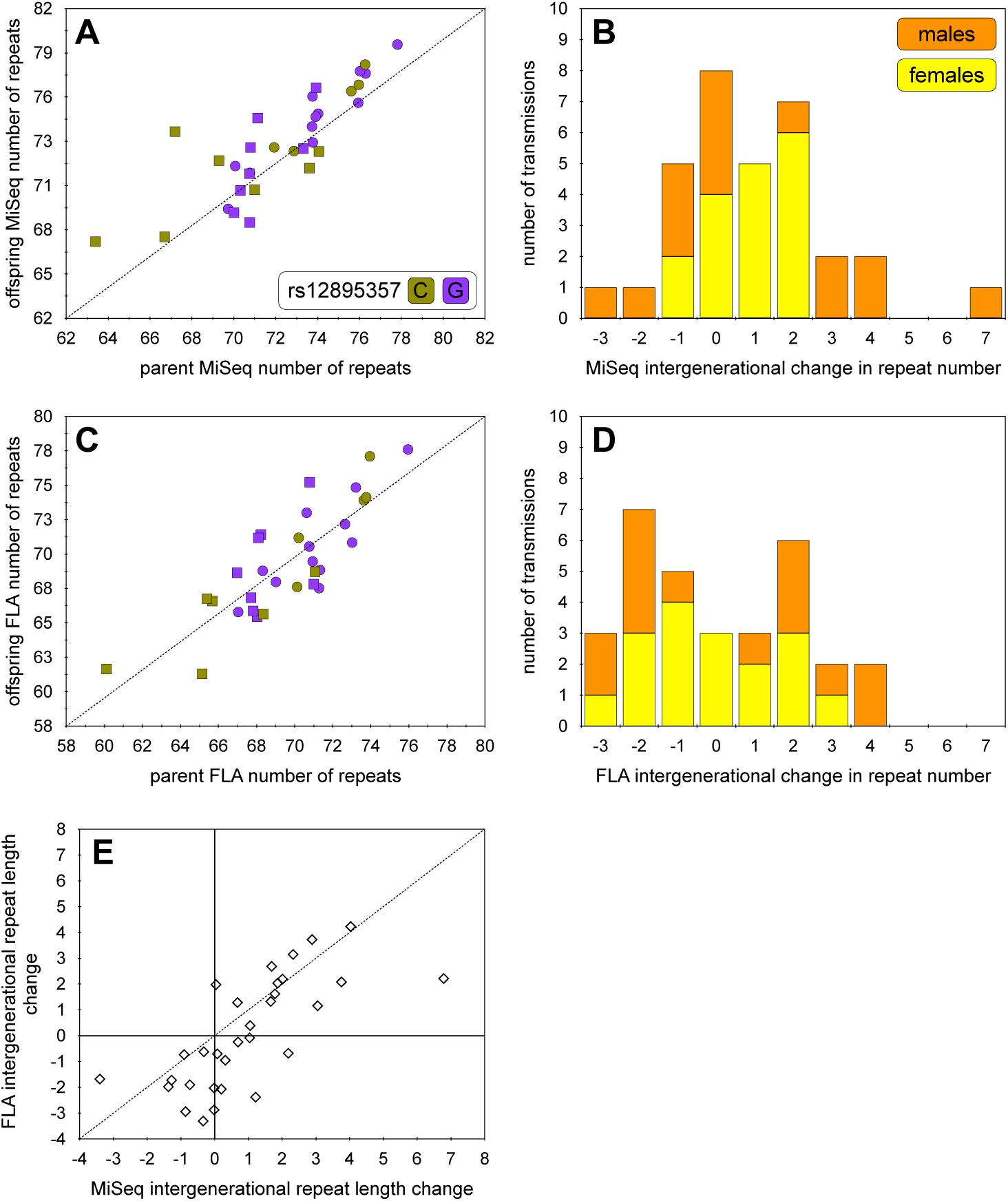
Intergenerational transmissions in SCA3. **A)** The scatterplot shows the association between *ATXN3* expanded disease-associated allele repeat length in parent and offspring in the Azorean SCA3 cohort as determined using the MiSeq analysis. The expected 1:1 line of no change is indicated (dashed line). **B)** The histogram shows the relative difference in *ATXN3* expanded disease-associated allele repeat length between parent and offspring in the Azorean SCA3 cohort for both paternal (orange) and maternal (yellow) transmissions as determined using the MiSeq analysis. **C)** The scatterplot shows the association between *ATXN3* expanded disease-associated allele repeat length in parent and offspring in the Azorean SCA3 cohort as determined using fragment length analysis (FLA). The expected 1:1 line of no change is indicated (dashed line). **D)** The histogram shows the relative difference in *ATXN3* expanded disease-associated allele repeat length between parent and offspring in the Azorean SCA3 cohort for both paternal (orange) and maternal (yellow) transmissions as determined using fragment length analysis (FLA). **E)** The scatterplot shows the association between *ATXN3* expanded disease-associated intergenerational transmissions allele as determined using fragment length (FLA) versus MiSeq analysis. The expected 1:1 line of methodological parity is indicated (dashed line). Note, that to allow visualisation of overlapping points, random jitter (up to +/- 0.4 repeats) has been applied to the number of repeats in scatterplots **A**, **C** and **E**.

**Figure S8.**
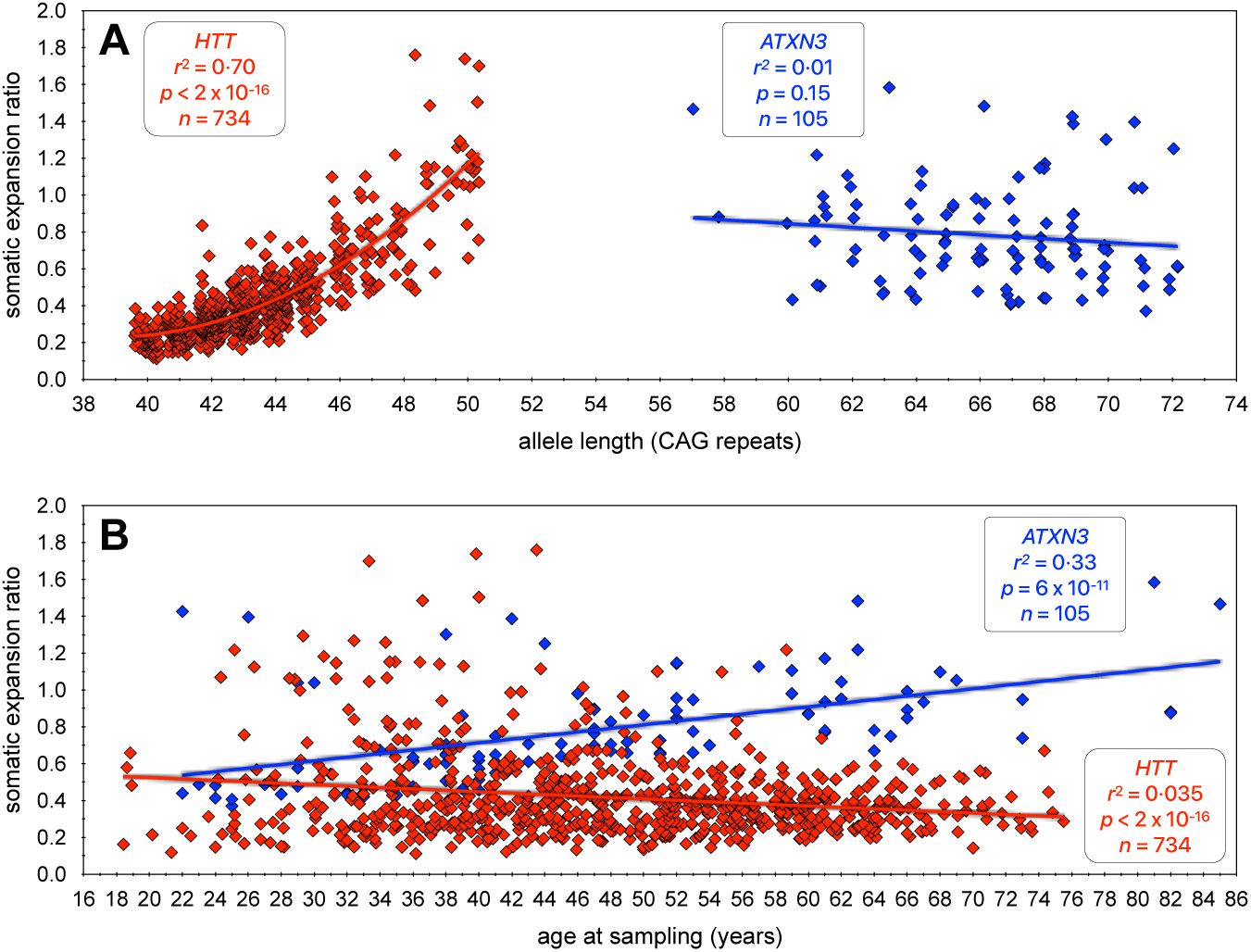
Contrast of the dynamics of somatic expansion of the *ATXN3* and *HTT* CAG repeats in the blood DNA. **A)** The scatterplot shows the ratio of somatic expansions of expanded disease-associated *ATXN3* (this study) and *HTT* (Ciosi *et al.,* (Ciosi *et al*. 2019)) alleles measured in blood DNA dependent on the number of pure CAG repeats. **B)** The scatterplot shows the ratio of somatic expansions of expanded disease-associated *ATXN3* (this study) and *HTT* (Ciosi *et al.,* (Ciosi *et al*. 2019)) alleles measured in blood DNA dependent on the age at sampling (years). Note, that to allow visualisation of overlapping points, random jitter (up to +/- 0.4 repeats) has been applied to the number of repeats.

## Notes

### Author Declarations

The Ethical Board of the University of the Azores (Parecer 1/2020) gave ethical approval for this work

## References

Afgan, E., Baker, D., van den Beek, M., Blankenberg, D., Bouvier, D., Cech, M., et al. 2016. The Galaxy platform for accessible, reproducible and collaborative biomedical analyses: 2016 update. Nucleic Acids Research 44: W3–W10. 10.1093/nar/gkw343

Akcimen, F., Martins, S., Liao, C., Bourassa, C.V., Catoire, H., Nicholson, G.A., et al. 2020. Genome-wide association study identifies genetic factors that modify age at onset in Machado-Joseph disease. Aging 12: 4742–4756. 10.18632/aging.102825

Auton, A., Abecasis, G.R., Altshuler, D.M., Durbin, R.M., Abecasis, G.R., Bentley, D.R., et al. 2015. A global reference for human genetic variation. Nature 526: 68–74. 10.1038/nature15393

Benn, C., Gibson, K., and Reynolds, D.D. 2021. Drugging DNA damage repair pathways for trinucleotide repeat expansion diseases. Journal of Huntington’s Disease 10: 203–220. 10.3233/JHD-200421

Brock, G.J.R., Anderson, N.H., and Monckton, D.G. 1999. *Cis*-acting modifiers of expanded CAG/CTG triplet repeat expandability: associations with flanking GC content and proximity to CpG islands. Human Molecular Genetics 8: 1061–1067. 10.1093/hmg/8.6.1061

Bunting, E.L., Hamilton, J., and Tabrizi, S.J. 2022. Polyglutamine diseases. Current Opinions in Neurobiology 72: 39–47. 10.1016/j.conb.2021.07.001

Cancel, G., Abbas, N., Stevanin, G., Dürr, A., Chneiweiss, H., Néri, C., et al. 1995. Marked phenotypic heterogeneity associated with expansion of a CAG repeat sequence at the spinocerebellar ataxia 3/Machado-Joseph disease locus. American Journal of Human Genetics 57: 809–816.

Cancel, G., Gourfinkel-An, I., Stevanin, G., Didierjean, O., Abbas, N., Hirsch, E., et al. 1998. Somatic mosaicism of the CAG repeat expansion in spinocerebellar ataxia type 3/Machado-Joseph disease. Human Mutation 11: 23–27. 10.1002/(SICI)1098-1004(1998)11:1<23::AID-HUMU4>3.0.CO;2-M

Ciosi, M., Cumming, S.A., Alshammari, A.M., Symeonidi, E., Herzyk, P., McGuinness, D., et al. 2018. Library preparation and MiSeq sequencing for the genotyping-by-sequencing of the Huntington disease *HTT* exon one trinucleotide repeat and the quantification of somatic mosaicism. Protocol Exchange. 10.1038/protex.2018.089

Ciosi, M., Maxwell, A., Cumming, S.A., Hensman Moss, D.J., Alshammari, A.M., Flower, M.D., et al. 2019. A genetic association study of glutamine-encoding DNA sequence structures, somatic CAG expansion, and DNA repair gene variants, with Huntington disease clinical outcomes. EBioMedicine 48: 568–580. 10.1016/j.ebiom.2019.09.020

de Mattos, E.P., Kolbe Musskopf, M., Bielefeldt Leotti, V., Saraiva-Pereira, M.L., and Jardim, L.B. 2019. Genetic risk factors for modulation of age at onset in Machado-Joseph disease/spinocerebellar ataxia type 3: a systematic review and meta-analysis. Journal of Neurology Neurosurgery and Psychiatry 90: 203–210. 10.1136/jnnp-2018-319200

Depienne, C., and Mandel, J.L. 2021. 30 years of repeat expansion disorders: What have we learned and what are the remaining challenges? American Journal of Human Genetics 108: 764–785. 10.1016/j.ajhg.2021.03.011

Durr, A., Stevanin, G., Cancel, G., Duyckaerts, C., Abbas, N., Didierjean, O., et al. 1996. Spinocerebellar ataxia 3 and Machado-Joseph disease: clinical, molecular, and neuropathological features. Annals Of Neurology 39: 490–499. 10.1002/ana.410390411

Gardiner, S.L., Boogaard, M.W., Trompet, S., de Mutsert, R., Rosendaal, F.R., Gussekloo, J., et al. 2019. Prevalence of Carriers of Intermediate and Pathological Polyglutamine Disease-Associated Alleles Among Large Population-Based Cohorts. JAMA Neurology 76: 650–656. 10.1001/jamaneurol.2019.0423

Genetic Modifiers of Huntington’s Disease Consortium, Lee, J.M., Correia, K., Loupe, J., Kim, K.-H., Barker, D., et al. 2019. CAG repeat not polyglutamine length determines timing of Huntington’s disease onset. Cell 178: 887–900. 10.1016/j.cell.2019.06.036

Grewal, R.P., Cancel, G., Leeflang, E.P., Durr, A., McPeek, M.S., Draghinas, D., et al. 1999. French Machado-Joseph disease patients do not exhibit gametic segregation distortion: a sperm typing analysis. Human Molecular Genetics 8: 1779–1784. 10.1093/hmg/8.9.1779

Hafford-Tear, N.J., Tsai, Y.C., Sadan, A.N., Sanchez-Pintado, B., Zarouchlioti, C., Maher, G.J., et al. 2019. CRISPR/Cas9-targeted enrichment and long-read sequencing of the Fuchs endothelial corneal dystrophy-associated TCF4 triplet repeat. Genetic Medicine 21: 2092–2102. 10.1038/s41436-019-0453-x

Hashida, H., Goto, J., Kurisaki, H., Mizusawa, H., and Kanazawa, I. 1997. Brain regional differences in the expansion of a CAG repeat in the spinocerebellar ataxias: Dentatorubral pallidoluysian atrophy, Machado Joseph disease, and spinocerebellar ataxia type 1. Annals Of Neurology 41: 505–511. 10.1002/ana.410410414

Hong, E.P., MacDonald, M.E., Wheeler, V.C., Jones, L., Holmans, P., Orth, M., et al. 2021. Huntington’s disease pathogenesis: two sequential components. Journal of Huntingtons Disease 10: 35–51. 10.3233/JHD-200427

Igarashi, S., Takiyama, Y., Cancel, G., Rogaeva, E.A., Sasaki, H., Wakisaka, A., et al. 1996. Intergenerational instability of the CAG repeat of the gene for Machado-Joseph disease (MJD1) is affected by the genotype of the normal chromosome: implications for the molecular mechanisms of the instability of the CAG repeat. Human Molecular Genetics 5: 923–932. 10.1093/hmg/5.7.923

Ito, Y., Tanaka, F., Yamamoto, M., Doyu, M., Nagamatsu, M., Riku, S., et al. 1998. Somatic mosaicism of the expanded CAG trinucleotide repeat in mRNAs for the responsible gene of Machado Joseph disease (MJD), dentatorubral pallidoluysian atrophy (DRPLA), and spinal and bulbar muscular atrophy (SBMA). Neurochemical Research 23: 25–32. 10.1023/a:1022441101801

Kawaguchi, Y., Okamoto, T., Taniwaki, M., Aizawa, M., Inoue, M., Katayama, S., et al. 1994. CAG expansions in a novel gene for Machado-Joseph disease at chromosome 14q32.1. Nature Genetics 8: 221–227. 10.1038/ng1194-221

Klockgether, T., Mariotti, C., and Paulson, H.L. 2019. Spinocerebellar ataxia. Nature Reviews of Disease Primers 5: 24. 10.1038/s41572-019-0074-3

Li, H., and Durbin, R. 2009. Fast and accurate short read alignment with Burrows–Wheeler transform. Bioinformatics 25: 1754–1760. 10.1093/bioinformatics/btp324

Li, H., and Durbin, R. 2010. Fast and accurate long-read alignment with Burrows–Wheeler transform. Bioinformatics 26: 589–595. 10.1093/bioinformatics/btp698

Li, H., Handsaker, B., Wysoker, A., Fennell, T., Ruan, J., Homer, N., et al. 2009. The Sequence Alignment/Map format and SAMtools. Bioinformatics 25: 2078–2079. 10.1093/bioinformatics/btp352

Lima, M., Mayer, F.M., Coutinho, P., and Abade, A. 1998. Origins of a mutation: population genetics of Machado-Joseph disease in the Azores (Portugal). Human Biology 70: 1011–1023.

Lima, M., Raposo, M., Ferreira, A., Melo, A.R.V., Pavao, S., Medeiros, F., et al. 2023. The Homogeneous Azorean Machado-Joseph Disease Cohort: Characterization and Contributions to Advances in Research. Biomedicines 11. 10.3390/biomedicines11020247

Lopes-Cendes, I., Maciel, P., Kish, S., Gaspar, C., Robitaille, Y., Clark, H.B., et al. 1996. Somatic mosaicism in the central nervous system in spinocerebellar ataxia type 1 and Machado-Joseph disease. Annals Of Neurology 40: 199–206. 10.1002/ana.410400211

Lysenko, L., Grewal, R.P., Ma, W., and Peddareddygari, L.R. 2010. Homozygous Machado Joseph Disease: a case report and review of literature. Canadian Journal of Neurological Sciences 37: 521–523. 10.1017/s0317167100010581

Maciel, P., Gaspar, C., DeStefano, A.L., Silveira, I., Coutinho, P., Radvany, J., et al. 1995. Correlation between CAG repeat length and clinical features in Machado-Joseph disease. American Journal of Human Genetics 57: 54–61.

Maciel, P., Gaspar, C., Guimaraes, L., Goto, J., Lopes-Cendes, I., Hayes, S., et al. 1999. Study of three intragenic polymorphisms in the Machado-Joseph disease gene (MJD1) in relation to genetic instability of the (CAG)n tract. European Journal of Human Genetics 7: 147–156. 10.1038/sj.ejhg.5200264

Maciel, P., Lopes-Cendes, I., Kish, S., Sequeiros, J., and Rouleau, G.A. 1997. Mosaicism of the CAG repeat in CNS tissue in relation to age at death in spinocerebellar ataxia type 1 and Machado-Joseph disease patients. American Journal of Human Genetics 60: 993–996.

Martin, M. 2011. Cutadapt removes adapter sequences from high-throughput sequencing reads. EMBnet.journal 17: 10–12. 10.14806/ej.17.1.200

Martins, S., Coutinho, P., Silveira, I., Giunti, P., Jardim, L.B., Calafell, F., et al. 2008. Cis-acting factors promoting the CAG intergenerational instability in Machado-Joseph disease. American Journal of Medical Genetics. Part B, Neuropsychiatric Genetics 147B: 439-446. 10.1002/ajmg.b.30624

Mätlik, K., Baffuto, M., Kus, L., Deshmukh, A.L., Davis, D.A., Paul, M.R., et al. 2024. Cell-type-specific CAG repeat expansions and toxicity of mutant Huntingtin in human striatum and cerebellum. Nature Genetics. 10.1038/s41588-024-01653-6

Mergener, R., Furtado, G.V., de Mattos, E.P., Leotti, V.B., Jardim, L.B., and Saraiva-Pereira, M.L. 2020. Variation in DNA Repair System Gene as an Additional Modifier of Age at Onset in Spinocerebellar Ataxia Type 3/Machado-Joseph Disease. Neuromolecular Medicine 22: 133–138. 10.1007/s12017-019-08572-4

Milne, I., Stephen, G., Bayer, M., Cock, P.J., Pritchard, L., Cardle, L., et al. 2013. Using Tablet for visual exploration of second-generation sequencing data. Briefings in Bioinformatics 14: 193–202. 10.1093/bib/bbs012

Munoz, E., Rey, M.J., Mila, M., Cardozo, A., Ribalta, T., Tolosa, E., and Ferrer, I. 2002. Intranuclear inclusions, neuronal loss and CAG mosaicism in two patients with Machado-Joseph disease. Journal of Neurological Sciences 200: 19–25. 10.1016/s0022-510x(02)00110-7

Nakano, K.K., Dawson, D.M., and Spence, A. 1972. Machado disease. A hereditary ataxia in Portuguese emigrants to Massachusetts. Neurology 22: 49–55. 10.1212/wnl.22.1.49

Nestor, C.E., and Monckton, D.G. 2011. Correlation of inter-locus polyglutamine toxicity with CAG•CTG triplet repeat expandability and flanking genomic DNA GC content. PLoS One 6: e28260. 10.1371/journal.pone.0028260

Ramos, A., Raposo, M., Mila, M., Bettencourt, C., Houlden, H., Cisneros, B., et al. 2016. Verification of Inter-laboratorial Genotyping Consistency in the Molecular Diagnosis of Polyglutamine Spinocerebellar Ataxias. Journal of Molecular Neuroscience 58: 83–87. 10.1007/s12031-015-0646-y

Raposo, M., Bettencourt, C., Melo, A.R.V., Ferreira, A.F., Alonso, I., Silva, P., et al. 2022. Novel Machado-Joseph disease-modifying genes and pathways identified by whole-exome sequencing. Neurobiology of Disease 162: 105578. 10.1016/j.nbd.2021.105578

Sasaki, H., Wakisaka, A., Fukazawa, T., Iwabuchi, K., Hamada, T., Takada, A., et al. 1995. CAG repeat expansion of Machado-Joseph disease in the Japanese: analysis of the repeat instability for parental transmission, and correlation with disease phenotype. Journal of Neurological Sciences 133: 128–133. 10.1016/0022-510x(95)00175-2

Scott, S.S.O., Pedroso, J.L., Barsottini, O.G.P., Franca-Junior, M.C., and Braga-Neto, P. 2020. Natural history and epidemiology of the spinocerebellar ataxias: Insights from the first description to nowadays. Journal of Neurological Sciences 417: 117082. 10.1016/j.jns.2020.117082

Sequeiros, J., Martindale, J., Seneca, S., Giunti, P., Kamarainen, O., Volpini, V., et al. 2010a. EMQN Best Practice Guidelines for molecular genetic testing of SCAs. European Journal of Human Genetics 18: 1173–1176. 10.1038/ejhg.2010.8

Sequeiros, J., Seneca, S., and Martindale, J. 2010b. Consensus and controversies in best practices for molecular genetic testing of spinocerebellar ataxias. European Journal of Human Genetics 18: 1188–1195. 10.1038/ejhg.2010.10

Shinde, D., Lai, Y., Sun, F., and Arnheim, N. 2003. Taq DNA polymerase slippage mutation rates measured by PCR and quasi-likelihood analysis: (CA/GT)n and (A/T)n microsatellites. Nucleic Acids Research 31: 974–980. 10.1093/nar/gkg178

Souza, G.N., Kersting, N., Krum-Santos, A.C., Santos, A.S., Furtado, G.V., Pacheco, D., et al. 2016. Spinocerebellar ataxia type 3/Machado-Joseph disease: segregation patterns and factors influencing instability of expanded CAG transmissions. Clinical Genetics 90: 134–140. 10.1111/cge.12719

Takiyama, Y., Sakoe, K., Soutome, M., Namekawa, M., Ogawa, T., Nakano, I., et al. 1997. Single sperm analysis of the CAG repeats in the gene for Machado-Joseph disease (MJD1): evidence for non-Mendelian transmission of the MJD1 gene and for the effect of the intragenic CGG/GGG polymorphism on the intergenerational instability. Human Molecular Genetics 6: 1063–1068. 10.1093/hmg/6.7.1063

Tanaka, F., Sobue, G., Doyu, M., Ito, Y., Yamamoto, M., Shimada, N., et al. 1996. Differential pattern in tissue specific somatic mosaicism of expanded CAG trinucleotide repeat in dentatorubral pallidoluysian atrophy, Machado Joseph disease, and X linked recessive spinal and bulbar muscular atrophy. Journal of Neurological Sciences 135: 43–50. 10.1016/0022-510x(95)00249-2

The R Core Team. 2016. “R: a language and environment for statistical computing.” In. Vienna, Austria: R Foundation for Statistical Computing.

The RStudio Team. 2016. “RStudio: integrated development for R.” In. Boston, MA: RStudio, Inc.

Wheeler, V.C., and Dion, V. 2021. Modifiers of CAG/CTG Repeat Instability: Insights from Mammalian Models. Journal of Huntington’s Disease 10: 123–148. 10.3233/JHD-200426

Woerner, A.E., Mandape, S., King, J.L., Muenzler, M., Crysup, B., and Budowle, B. 2021. Reducing noise and stutter in short tandem repeat loci with unique molecular identifiers. Forensic Science International: Genetics 51. 10.1016/j.fsigen.2020.102459

Woods, B.T., and Schaumburg, H.H. 1972. Nigro-spino-dentatal degeneration with nuclear ophthalmoplegia. A unique and partially treatable clinico-pathological entity. Journal of Neurological Sciences 17: 149–166. 10.1016/0022-510x(72)90137-2

Wright, G.E.B., Collins, J.A., Kay, C., McDonald, C., Dolzhenko, E., Xia, Q., et al. 2019. Length of uninterrupted CAG, independent of polyglutamine size, results in increased somatic instability, hastening onset of Huntington disease. American Journal of Human Genetics 104: 1116–1126. 10.1016/j.ajhg.2019.04.007

